# Recurrent stroke prediction by applying a stroke polygenic risk score in the Japanese population

**DOI:** 10.1101/2024.06.17.24309034

**Authors:** Naoki Kojima, Masaru Koido, Yunye He, Yuka Shimmori, Tsuyoshi Hachiya, BioBank Japan, Stéphanie Debette, Yoichiro Kamatani

## Abstract

**Background:** Recently, various polygenic risk score (PRS)-based methods were developed to improve stroke prediction. However, current PRSs (including cross-ancestry PRS) poorly predict recurrent stroke. Here, we aimed to determine whether the best PRS for Japanese individuals can also predict stroke recurrence in this population by extensively comparing the methods and maximizing the predictive performance for stroke onset.

**Methods:** We used data from the BioBank Japan (BBJ) 1^st^ cohort (n=179,938) to derive and optimize the PRSs using a 10-fold cross-validation. We integrated the optimized PRSs for multiple traits, such as vascular risk factors and stroke subtypes to generate a single PRS using the meta-scoring approach (metaGRS). We used an independent BBJ 2^nd^ cohort (n=41,929) as a test sample to evaluate the association of the metaGRS with stroke and recurrent stroke.

**Results:** We analyzed recurrent stroke cases (n=174) and non-recurrent stroke controls (n=1,153) among subjects within the BBJ 2^nd^ cohort. After adjusting for known risk factors, metaGRS was associated with stroke recurrence (adjusted OR per SD 1.18 [95% CI: 1.00–1.39, p=0.044]), although no significant correlation was observed with the published PRSs. We administered three distinct tests to consider the potential index event bias; however, the outcomes derived from these examinations did not provide any significant indication of the influence of index event bias. The high metaGRS group without a history of hypertension had a higher risk of stroke recurrence than that of the low metaGRS group (adjusted OR 2.24 [95% CI: 1.07–4.66, p=0.032]). However, this association was weak in the hypertension group (adjusted OR 1.21 [95% CI: 0.69–2.13, p=0.50]).

**Conclusions:** The metaGRS developed in a Japanese cohort predicted stroke recurrence in an independent cohort of patients. In particular, it predicted an increased risk of recurrence among stroke patients without hypertension. These findings provide clues for additional genetic risk stratification and help in developing personalized strategies for stroke recurrence prevention.

## Introduction

Stroke is a major cause of mortality in Japan, with 56,000 deaths reported in 2020.^1^ The conventional risk factors for stroke include hypertension, high waist-to-hip ratio, smoking, cardiac causes, dyslipidemia, and diabetes mellitus.^2^ In Japan, the stroke recurrence rate is up to 30–50% during 5-10 years of follow-up after the first stroke.^3,4^ Accordingly, it will be medically beneficial to stratify high-risk groups for recurrent stroke among those who have experienced a stroke to potentially generate more intensive secondary prevention strategies than current recommendations.

Genome-wide association studies (GWAS) have identified many disease-susceptibility variants associated with complex traits.^5^ A polygenic risk score (PRS) is the weighted summation of the individual genetic effects of these variants. Its weighting strategy varies depending on the construction method; traditionally, only significant variants are used in developing this score. The recently developed PRS methods involve non-significant variants and updated effect weights and consider the linkage disequilibrium (LD) structure. The development of PRS methods has helped stratify high-risk groups for complex traits,^6–11^ including stroke.^12^

Polygenic risk scores developed using the 32 genome-wide significant (p< 5.0×10^-8^) variants or 90 marginally associated (p<1.0×10^-5^) variants (PRS90) from the MEGASTROKE study^12^ are associated with stroke onset in subjects of European ancestry.^12,13^ The meta-scoring PRS approach (metaGRS) includes 3.2 million variants by combining PRSs for stroke subtypes, risk factors, and comorbidities by adjusting the effect weight via elastic-net logistic regression; this approach has an improved predictive performance for stroke compared to that of PRS90.^14^ MetaGRS can predict stroke incidence independent of environmental factors and could help motivate individuals with high genetic risk to make lifestyle changes for stroke prevention (although not yet implemented in clinical practice outside a research setting).^15^ The PRS shows reduced transferability between populations. Additionally, a PRS developed using various variants derived from Japanese GWAS successfully predicted stroke onset in the Japanese population.^16,17^ Most recently, the GIGASTROKE study proposed an integrated PRS approach among PRSs derived from populations of multiple ancestries using the metaGRS framework (iPGS), which showed a better predictive ability than the MEGASTROKE European or East Asian PRS.^18^ However, the PRS did not successfully predict stroke recurrence; for example, PRS32 and iPGS did not significantly predict stroke recurrence after adjusting for clinical comorbidities, with notably smaller effect sizes than for non-recurrent stroke.^12,18^ Furthermore, the potential effect of index event (also known as “collider”) bias that may distort the association of PRS was suspected.^19,20^

The optimal method to improve the predictive accuracy of PRS depends on the population- and trait-specific genetic architecture.^21–30^ Therefore, we compared different PRS methods and determined whether the best PRS can predict the onset of recurrent stroke in a Japanese population.

## Methods

The workflow of this study is shown in Figure 1. This article follows the TRIPOD (Transparent reporting of a multivariable prediction model for individual prognosis or diagnosis) reporting guidelines.

**Figure 1.**
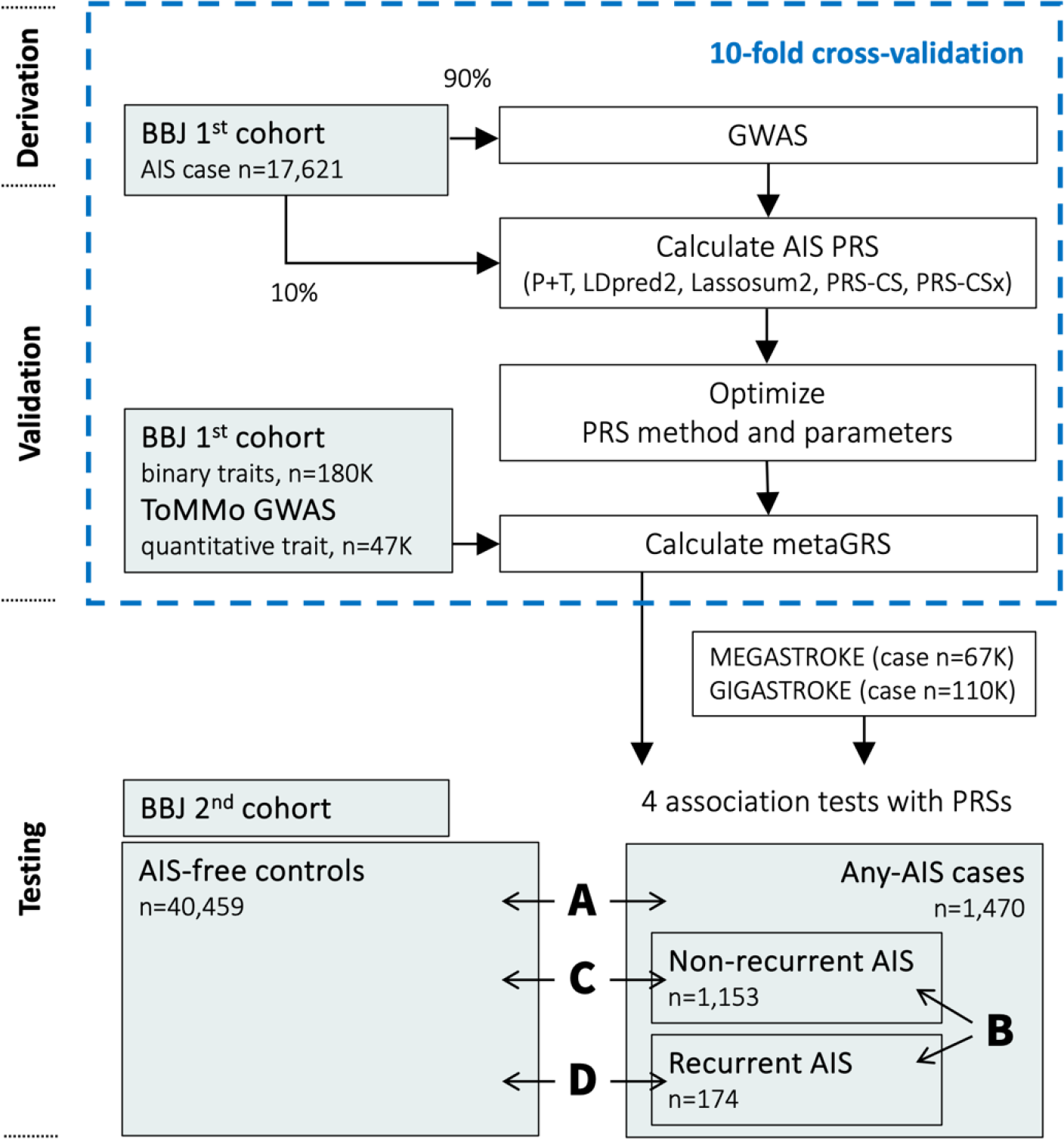
Workflow. MEGASTROKE AIS summary statistics of European (EUR) studies were only used for PRS-CSx. 1000 Genomes Project super population samples (EAS or EUR) were used for the LD reference panel. Abbreviations: GWAS = genome-wide association study, PRS = polygenic risk score, P+T = pruning and thresholding, OR = odds ratio, AIS = all ischemic stroke, ToMMo = Tohoku Medical Megabank; LD, linkage disequilibrium. We used a logistic regression model to assess the association of the PRS using the two case control settings for AIS (A: any-AIS vs. AIS-free controls) and AIS recurrence (B: recurrent AIS vs. non-recurrent AIS). We also applied two other combinations of case-controls (C: recurrent versus AIS-free controls and D: non-recurrent versus AIS-free controls).

### Study subjects and quality control

BioBank Japan (BBJ) involves physicians diagnosing all ischemic stroke (AIS) cases at the collaborating hospitals. BBJ was established in 2003 and recruited 267,000 patients from 12 medical institutions (66 hospitals) in two phases.^31–33^ The recruited patients had at least one of the 51 primarily multifactorial (common) diseases, which accounted for 440,000 cases. We used BBJ 1^st^ cohort (BBJ1) data to derive PRSs and available independent BBJ 2^nd^ cohort (BBJ2) data to evaluate the performance of PRSs in predicting AIS and recurrent AIS. Recurrent AIS information was unavailable for the BBJ1 data. In BBJ2, any AIS cases (n=1,470), AIS-free controls (n=40,459), recurrent AIS cases (n=174), and non-recurrent AIS controls (n=1,153) were available. The mean duration from the first episode of AIS onset to recurrent AIS onset was 4.88 years. Detailed sample characteristics are listed in Supplementary Methods and Table 1.

**Table 1.**
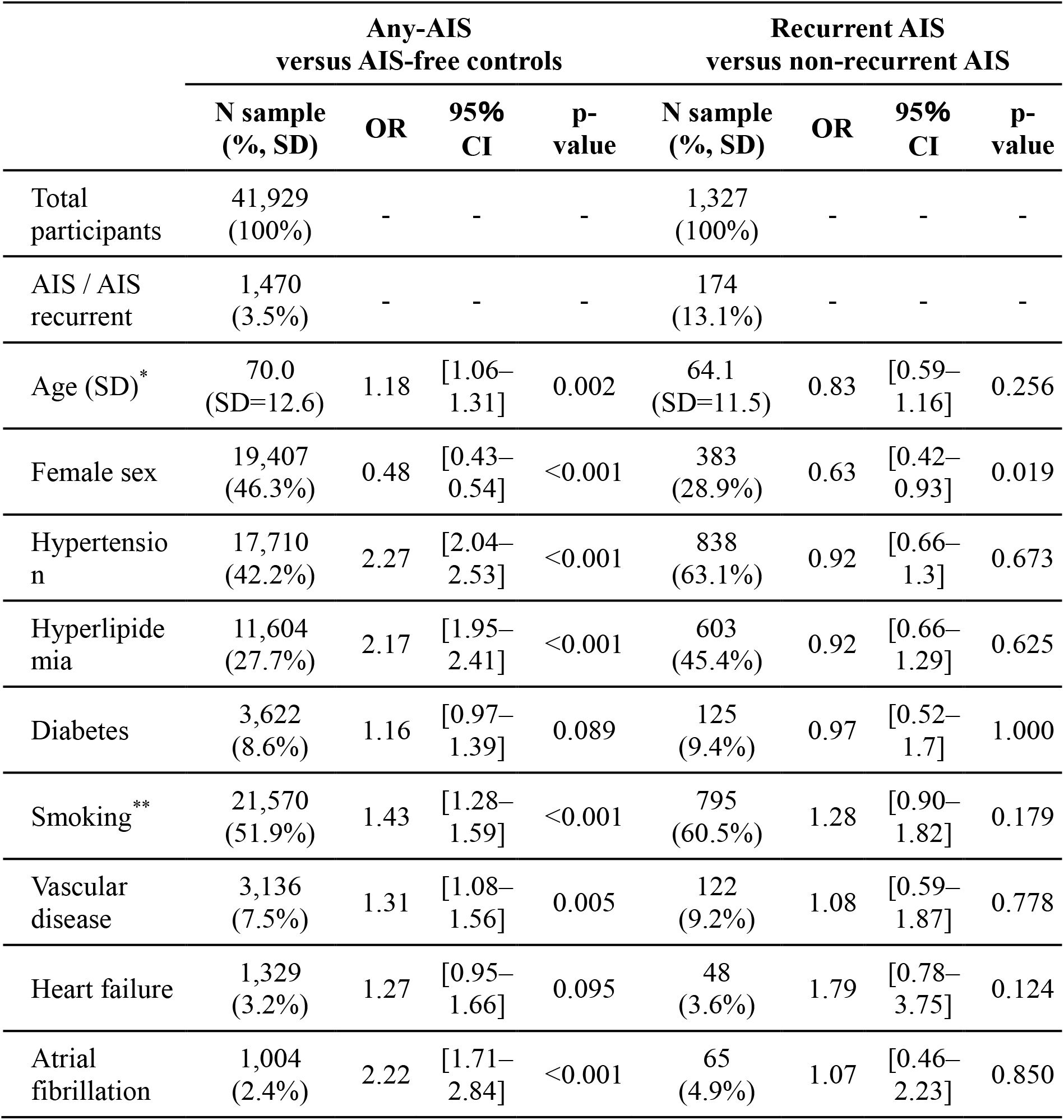
Characteristic of the testing sample (BBJ2) All risk factor characteristics were derived from history and not at the time of registration. Odds ratio (OR), 95% CI (95% confidence intervals, and p-values were calculated using logistic regression for onset and AIS recurrence (unadjusted for other factors). *Age at AIS case-control was that at recruitment, while age at recurrent AIS case-control was that at first incidence. The numbers indicate the median threshold age. ** The total number of missing values of smoking was 389 for all case-control and 12 for recurrent AIS case-control samples. Abbreviations: BBJ2, BioBank 2^nd^ cohort; AIS, all ischemic stroke; SD = standard deviation

This study was approved by the ethics committee of the Institute of Medical Science, the University of Tokyo, Japan. Quality control, pre-phasing, and genotype imputation were conducted using PLINK(v2.0),^34–37^ Eagle (v2.4.1), and Minimac4 (v1.0.2), respectively. The detailed processes are presented in the Supplementary Methods.

### Constructing PRSs for AIS

Unbiased PRSs were obtained by applying a 10-fold cross-validation to select the model and optimize the parameters.^26,38^ Briefly, BBJ1 samples were randomly split into ten equal-sized subsamples. We retained one subsample for validation and the others for training. We repeated this process 10 times, with each of the ten subsamples used exactly once for validation. A GWAS was conducted on the training set in each iteration, adjusted for age, sex, and the first 10 principal components (PCs) via Firth logistic regression using PLINK (v.2.0).^34^

We obtained the weights of variants for PRS from the GWAS summary statistics of the training set using five PRS methods—P+T (PLINK (v.1.9)^35^ for clumping), LDpred2,^39^ Lassosum2,^40^ (LDpred2 and Lassosum2 by bigsnpr package (v.1.7.2) in R (v.3.5.0)), PRS-CS (v.1.0.0),^41^ and PRS-CSx (v.1.0.0).^42^ The PRS-CSx integrated BBJ1 with the European stroke GWAS summary statistics (MEGASTROKE; the largest study available at this study design)^43^ by learning an optimal linear combination. We used combinations of parameters for P+T (1,224 parameters), LDpred2 (126 parameters), Lassosum2 (200 parameters), PRS-CS (9 parameters), and PRS-CSx (9 parameters), as described in the Supplementary Methods.

Subsequently, the PRSs for the validation sample were calculated using the weights obtained from the training samples. The accuracy for predicting AIS cases was evaluated from Nagelkerke’s R^2^ (simply “R^2^” from this point onwards)^29,44^ after adjusting for age, sex, and the first 10 PCs. We calculated the mean R^2^ over 10 cross-validation results for each method with each parameter after a 10-fold cross-validation. We chose the method and parameters that maximized incremental R^2^ (PRSAIS) among these PRSs.

We further integrated the PRSAIS with PRSs of vascular risk factors, such as stroke subtypes and comorbid diseases presence, using the elastic net framework to construct a metaGRS using the glmnet package (v.4.1.3) in R (v.4.1.0). Nine binary traits and eight quantitative traits of vascular risk factors reported in a previous study^14^ are described in the Supplementary Methods. Binary traits were determined by conducting GWAS and attempting to obtain unbiased weights using cross-validations. The effect weights from the derivation sample every 17 traits were calculated using PRS-CS-auto since it did not require an independent validation sample set for parameter optimization and performed well for various traits.^21,26,39,41,45^ Subsequently, we used a validation sample to calculate the weight of AIS and the 17-trait PRSs to predict AIS using elastic-net logistic regression. We conducted a 10-fold cross-validation and used the mean weight for testing.

We used PLINK (v2.0)^34^ to calculate the individual PRS by aggregating the effect estimates multiplied by each imputed dosage into a single score per person.

### Risk factors

The following seven risk factors that were previously utilized as covariates^12^ were used as covariates for testing: hypertension (systolic blood pressure>140 mmHg, diastolic blood pressure>90 mmHg, or hypertension history), hyperlipidemia, diabetes (all types), smoking (current smoker), vascular disease (myocardial infarction, peripheral artery disease, stable angina pectoris, and unstable angina pectoris), congestive heart failure, and atrial fibrillation (including atrial flutter). A sample was considered to have a risk factor status if it had that status at enrollment or from historical records (Tables 1 and S1).

### Assessment of the association of PRS with AIS and AIS recurrence

We used a single selected method with optimized parameters and calculated the metaGRS in independent testing sample sets. We used a logistic regression model to assess the association of the PRS using the two case-control settings for AIS (any-AIS versus AIS-free controls) and AIS recurrence (recurrent AIS versus non-recurrent AIS). We also applied two other combinations of case-controls: recurrent AIS versus AIS-free controls and non-recurrent AIS versus AIS-free controls (Figure 1). Furthermore, we examined additional PRS contributions of the seven risk factors to predictive accuracy and discriminative performance using the values of R^2^ and area under the curve (AUC), according to our previous studies.^14,46^ Additionally, we evaluated the performance of the following PRSs derived from other milestone studies for stroke prediction: 32 genome-wide significant variants (for any stroke, ischemic stroke, or ischemic stroke subtypes) from the MEGASTROKE cross-ancestry study of 524,354 individuals (PRS32),^43^ 89 genome-wide significant variants of 1,614,080 multi-population individuals (PRS89), and 6,010,730 variants of the East Asian PRS developed from 9,809 individuals (iPGSEAS),^18^ both from the GIGASTROKE study. The PRS calculation process is described in the Supplementary Methods.

### Considering potential index event bias

An index event bias may be induced when the samples are only selected from cases.^47^ We evaluated the extent to which the index event bias affected our results since we used case-only samples in this study. The association of PRSs with recurrent AIS was evaluated after adjusting for seven risk factors, in addition to age, sex, and the first 10 PCs. Adjusting for such confounding bias will not be enough to eliminate bias for a recurrence association study.^48^ Therefore, we sought to mitigate a potential index event bias by applying three distinct methodologies.^48,49^ First, we utilized linear and logistic regression models to assess the relationships between metaGRS and covariates within the any-AIS case (n=1,327) and AIS-free control groups (n=40,459). Initially, we did not adjust for age, sex, the seven risk factors, or the first 10 PCs. Subsequently, we observed the distributions of covariate values across the metaGRS quintiles. We performed statistical tests to detect heterogeneity in the estimates between the prevalent case and control groups, following the methodology of a prior study.^19,50^ Second, we refined our analysis by adjusting for associations between metaGRS and AIS recurrence while considering the differential effects of covariates, according to a previous method.^19^ Finally, we applied the inverse probability weighted (IPW) approach^47^ to comprehensively account for index event bias. Collectively, these analytical approaches were adopted to enhance the validity of our findings.

### Association of metaGRS and AIS recurrence in patients with/without hypertension

Logistic regression was conducted in subgroups with and without hypertension and metaGRS tertiles among recurrent AIS cases (with hypertension n=107, without hypertension n=67) and non-recurrent AIS controls (with hypertension n=731, without hypertension n=422) to assess the relationship between the PRS and the risk of AIS recurrence. The low metaGRS tertile was set as the reference group and adjusted for age, sex, and the first 10 PCs.

### Statistical analysis

The mean with standard deviation (SD) or proportion of factors was reported for the baseline characteristics of testing samples. The incremental value (R^2^ or AUC) was estimated from the differences between patients with and without PRSs of the fitted values of age, sex, first 10 PCs, and seven risk factors^48,57,58^ and calculated as the 95% confidence interval. The pROC package (v.1.18.0) in R was used to determine the discriminative ability of the AUC. The IPW package (v.1.2) in R was used for the IPW approach. R (v. 3.5.0) was used to perform logistic regression to calculate R^2^, Pearson’s correlation coefficient, and linear regression. All statistical tests were two-sided. The significance level was set at p = 0.05.

## Results

### Derivation of effect weight

The imputed genotype data of 17,621 AIS cases and 162,317 controls without an AIS diagnosis were used for 9,622,629 autosomal variants after implementing quality control of the BBJ1 dataset (Tables S2-S4).

We conducted a 10-fold cross-validation to adjust the parameters and select the best PRS associated with AIS. We performed GWAS 10 times on 90% of the randomly selected BBJ1 dataset (training data). We successfully detected previously reported^43,51^ signals in each dataset (p<5×10^-8^), including *SH3PXD2A*, *CCDC63* (eight times), *CUX2*, and *LINC02356* (every time) (Figure S1, Table S5).

We confirmed some expected characteristics of each PRS method (such as low accuracy) using only genome-wide significant variants (Tables S6–10 and Supplementary Notes). The mean incremental R^2^ values of each scoring method with the best-performed parameters were 0.0038 (95% CI: 0.0030–0.0046), 0.00443 (95% CI: 0.0035–0.0054), 0.0039 (95% CI: 0.0030–0.0048), 0.00441 (95% CI: 0.0036–0.0053), and 0.0037 (95% CI: 0.0031–0.0042) for P+T, LDpred2, Lassosum2, PRS-CS, and PRS-CSx, respectively (Table 2). We chose LDpred2 with the parameter set of ρ-value = 0.0056, a heritability-value = 1.0×h^2^LDSC, where h^2^LDSC is the heritability estimate from the constrained LD score regression^52^, and a no-sparse model for subsequent analyses, since it showed the best mean incremental R^2^ value among the five methods.

**Table 2.** Polygenic risk score performance at validation.

| Method | Best parameters | Mean number of variants | Best mean incremental Nagelkerke $R^2$ | Standard deviation | 95% confidence interval |
| --- | --- | --- | --- | --- | --- |
| P+T | Clumping $R^2=0.95$,<br>Clumping kb=526,<br>Imputation $R^2=0.8$,<br>p-value threshold=1 | 3,144,737 | 0.0038 | 0.0012 | 0.0030–0.0046 |
| LDpred2 | $\rho$ value=0.0056,<br>heritability value x<br>1.0,<br>no sparse | 898,456 | 0.00443 | 0.0013 | 0.0035–0.0054 |
| Lassosum2 | S=0.9,<br>lambda=0.00388 | 282,520 | 0.0039 | 0.0013 | 0.0030–0.0048 |
| PRS-CS | Phi=1.00E-04 | 985,439 | 0.00441 | 0.0012 | 0.0036–0.0053 |
| PRS-CSx | Phi=1.00E-05 | 1,016,745 | 0.0037 | 0.0008 | 0.0031–0.0042 |

We observed an average number of nonzero weights for 8.4 traits after computing the metaGRS via elastic net regularization 10 times (10-fold). The metaGRS weight of AIS was highest (mean=0.123, SD=0.026), followed by diastolic blood pressure (mean=0.039, SD=0.039), atrial fibrillation (mean=0.023, SD=0.024), and myocardial infarction (mean=0.018, SD=0.025) (Figure S2 and Table S11). Only the triglyceride weights were zero at all 10 measurements among the 18 traits included in the metaGRS calculation. The number of variants used for metaGRS was 1,014,026; a total of 1,011,847 variants (99.8%) remained after matching with the BBJ2 dataset.

### Association of metaGRS with AIS cases and recurrent AIS

We used the imputed genotype data of 1,470 AIS cases and 40,459 controls without a diagnosis of AIS for 59,387,070 variants from the BBJ2 dataset to test the association of metaGRS with AIS and AIS recurrence. The AIS case-only sample of the BBJ2 was used to analyze AIS recurrence. Table 1 presents the characteristics of the test samples.

MetaGRS was associated with AIS diagnosis after adjusting for age, sex, first 10 PCs, and seven risk factors (adjusted OR, 1.21 [95% CI: 1.15–1.27, p=2.89×10^-12^]), as previously reported.^14,18^ MetaGRS was also associated with AIS recurrence compared with recurrence-free AIS (adjusted OR 1.18 [95% CI: 1.00–1.39, p=0.044]; Table 3 and Figure S3). MetaGRS showed stronger association when comparing recurrent AIS with AIS-free controls (adjusted OR 1.37 [95% CI: 1.18–1.59, p=5.35×10^-5^]; Table S12).

**Table 3.**
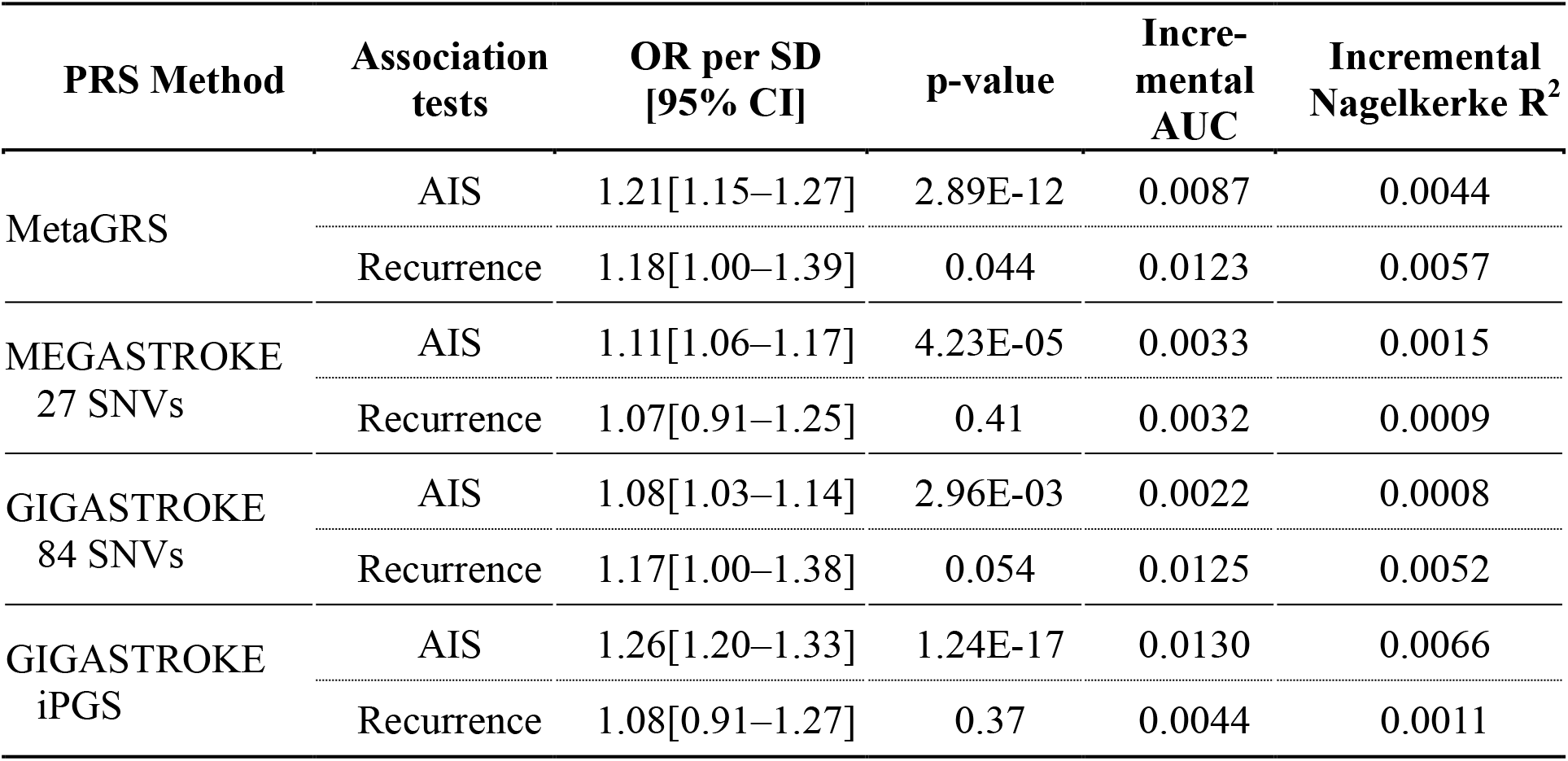
Polygenic risk score performance at testing. Polygenic risk score performance was evaluated using an independent testing set for AIS and recurrent AIS. We showed two main association tests; AIS (any-AIS cases vs. AIS-free controls) and AIS recurrence (recurrent AIS vs. non-recurrent AIS). Incremental AUC and R^2^ are the differences in the values when fitting with/without PRS, along with age, sex, the first 10 PCs, and seven risk factors. Abbreviations: AIS, all ischemic strokes; OR, odds ratio; AUC, area under the curve; PC, principal components; PRS, polygenic risk score; SD = standard deviation

| PRS Method | Association tests | OR per SD [95% CI] | p-value | Incremental AUC | Incremental Nagelkerke R<sup>2</sup> |
| --- | --- | --- | --- | --- | --- |
| MetaGRS | AIS | 1.21[1.15–1.27] | 2.89E-12 | 0.0087 | 0.0044 |
|  | Recurrence | 1.18[1.00–1.39] | 0.044 | 0.0123 | 0.0057 |
| MEGASTROKE<br>27 SNVs | AIS | 1.11[1.06–1.17] | 4.23E-05 | 0.0033 | 0.0015 |
|  | Recurrence | 1.07[0.91–1.25] | 0.41 | 0.0032 | 0.0009 |
| GIGASTROKE<br>84 SNVs | AIS | 1.08[1.03–1.14] | 2.96E-03 | 0.0022 | 0.0008 |
|  | Recurrence | 1.17[1.00–1.38] | 0.054 | 0.0125 | 0.0052 |
| GIGASTROKE<br>iPGS | AIS | 1.26[1.20–1.33] | 1.24E-17 | 0.0130 | 0.0066 |
|  | Recurrence | 1.08[0.91–1.27] | 0.37 | 0.0044 | 0.0011 |

The contribution of the metaGRS and traditional risk factors showed an AIS prediction accuracy with an R^2^ value of 0.06 and an AUC of 0.689 after constructing the baseline model using age, sex, the first 10 PCs, and seven risk factors. The incremental AUCs were 0.0087 and 0.0123 for AIS and AIS recurrence, respectively when metaGRS was added to the baseline model (Table 3). In our dataset, clinical risk factors (including hypertension) were related to AIS diagnosis but were insignificantly associated with AIS recurrence (Table S1). We assessed the prediction performance of previously developed PRSs for AIS and AIS recurrence in our dataset. After matching with the BBJ2 dataset (Supplementary Methods), 27, 84, and 5,756,652 variants remained in PRS32, PRS89, and iPGS, respectively. We confirmed their association with AIS diagnosis; adjusted ORs were 1.11 [95% CI: 1.06–1.17, p=4.23×10^-5^], 1.08 [95% CI: 1.03–1.14, p=2.96×10^-3^], and 1.26 [95% CI: 1.20–1.33, p=1.24×10^-17^] for PRS32, PRS89, and iPGS, respectively (Table 3, Figure S3); however, a significant association was not observed between PRSs and AIS recurrence (p-values of 0.41, 0.054, and 0.37, respectively; Table 3 and Figure S3). Our Japanese optimized metaGRS was the only PRS significantly associated with AIS recurrence in this study.

### Analyzing for potential index event bias

We observed the values of covariates at the AIS-free control group and any-AIS case group in each quintile. We did not find any significant heterogeneous relationships between the covariates and the metaGRS in terms of regression estimates in the prevalent case and control samples (p>0.05, Table S13).

We used three different variable models—i) age and sex; ii) age, sex, and seven risk factors; and iii) age, sex, the first 10 PCs, and seven risk factors—to determine the association between metaGRS and recurrent AIS; none of these confounders significantly influenced our results (Figure S4).

We compared the association results of IPW adjusted (accounting for index event bias) with those of non-adjusted IPW (accounting for confounding bias). The results remained almost unchanged, but the 95% confidence intervals overlapped (Figure S5). A comparison of the three distinct models did not indicate an effect of index event bias.

### Association of metaGRS and AIS recurrence in patients with/without hypertension

We divided the test sample into subgroups according to the presence or absence of a history of hypertension and evaluated the risk effect of the metaGRS tertile. The high metaGRS group without a history of hypertension showed a higher risk effect for AIS recurrence compared to the low metaGRS group (OR of the high metaGRS group compared to that of the low metaGRS group was 2.24 [95% CI: 1.07–4.66, p=0.032], Figure 2 and Table S14). However, no significant association was observed between the metaGRS and AIS recurrence in the group with a history of hypertension (the OR of the high metaGRS group compared to that of the low metaGRS group was 1.21 [95% CI: 0.69–2.13, p=0.50] (Figure 2 and Table S14).

**Figure 2.**
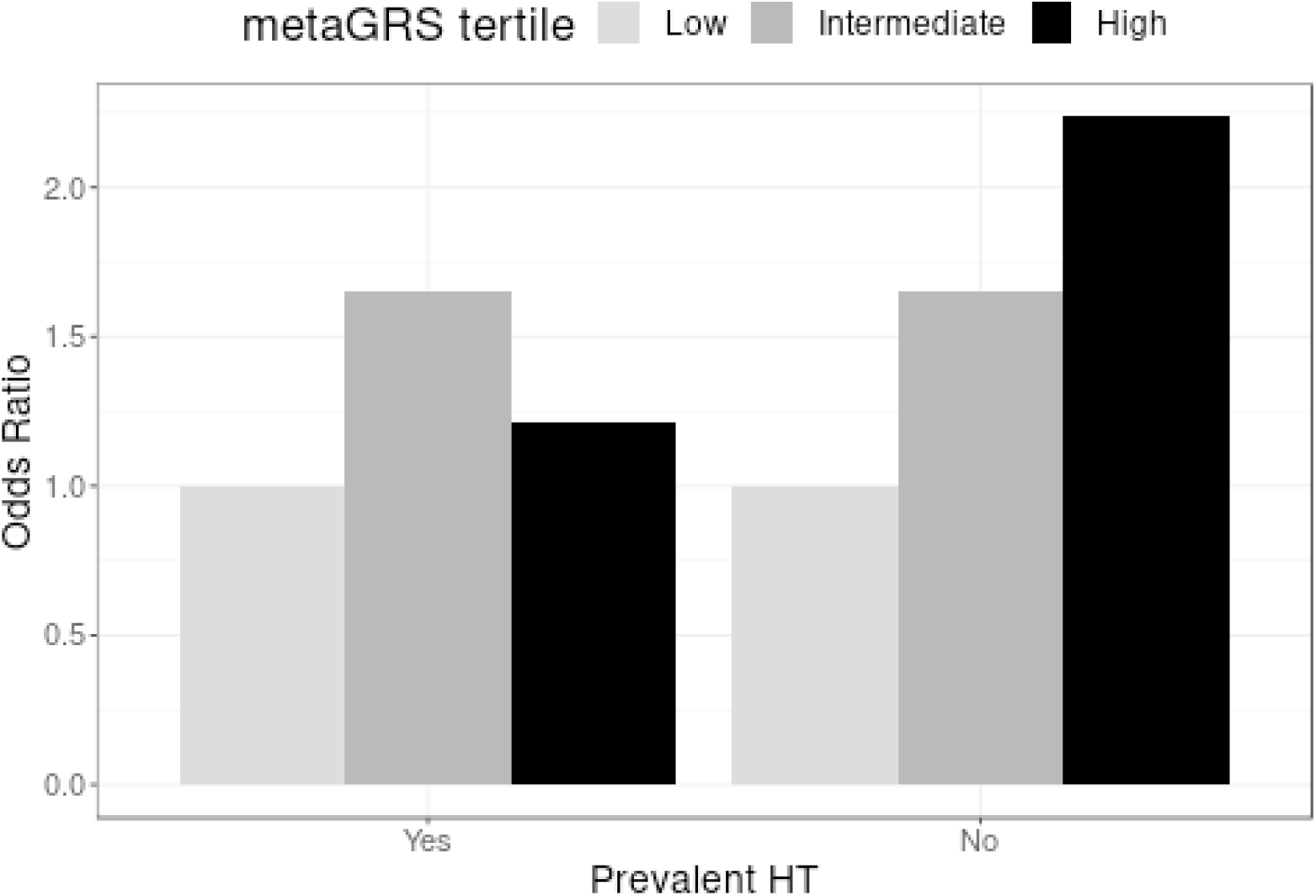
Odds ratio of metaGRS tertiles with/without a history of hypertension. Association of AIS recurrence and meta-GRS tertiles with or without a history of hypertension (HT) in the testing sample, with reference to the low metaGRS tertiles.

## Discussion

We successfully examined the association between recurrent AIS and our best model PRS (metaGRS using LDpred2); the adjusted OR was 1.18 for each unit of SD increase in PRS. Our metaGRS showed stronger (adjusted OR per SD=1.37) association when comparing recurrent AIS with AIS-free controls. Furthermore, a high PRS was associated with AIS recurrence particularly in groups without a history of hypertension (OR of the top vs. bottom metaGRS tertile=2.24). These results are consistent with the result of a previous study wherein the stroke prediction accuracy of the PRS was high in the group with low CHA2DS2-VASc scores.^12^ These results indicate the utility of the PRS in developing more precise strategies to prevent AIS recurrence in individuals with a high PRS who do not have high profiles based on clinical risk factors.

We attempted to mitigate potential index event bias since our purpose was to specifically determine the efficacy of PRS among AIS patients. It is difficult to predict and provide an accurate assessment of recurrent AIS based on genetic predisposition owing to the possible effect of index event bias leading to a distorted association in studies on recurrent stroke.^19,20,53^ This study found no evidence of heterogeneous associations between covariates and the metaGRS; we did not find any evidence of a solid collider bias of known variables. By applying IPW, we confirmed that our results support the association between metaGRS and recurrent AIS.

There are three putative reasons our metaGRS could predict AIS recurrence. First, the metaGRS algorithm combines the genetic profiles of related traits and slightly improves the performance, reaching the level of significance. Second, the performances of PRS-CS (incremental R^2^=0.00441) and LDpred2 (incremental R^2^=0.00443) in our validation analysis were better than those of other traditional PRS methods, such as P+T (incremental R^2^=0.0038) . This demonstrated the importance of using shrinkage estimation methods that consider LD to predict AIS and AIS recurrence. Third, we restricted to use only single matched ancestry throughout.

Nevertheless, our study had several limitations. First, the sample size for recurrent AIS needs to be increased (n=174 at testing), even in the largest hospital-based biobank in Japan. Compared to our metaGRS, iPGS constructed in GIGASTROKE showed a stronger association for AIS and weaker association for AIS recurrence. Although potential discrepancies exist, both PRSs (metaGRS and iPGS) exhibit the same direction of effects and have overlapping confidence intervals (Table 3, Figure S3). Second, despite using as many covariates (age, sex, the first 10 PCs, and seven risk factors (hypertension, hyperlipidemia, diabetes mellitus, smoking, vascular disease, congestive heart failure, and atrial fibrillation)) as possible based on a previous study,^12^ other confounders might have affected our results. Finally, there may have been an index event bias that was not fully detected by each method that we implemented; however this risk was minimized using multiple approaches. Further studies using different sample sets (including other ancestry groups) are warranted to confirm the prediction of recurrent stroke using the PRS.

In conclusion, our study indicated that PRS can be applied to predict AIS recurrence in addition to traditional clinical risk factors. This shows the potential utility of PRS in population-based screening and in the clinical setting. Overall, our results indicate that stratifying high-risk groups for recurrent stroke among those who have experienced a stroke could be medically beneficial and help in developing personalized strategies for recurrence prevention. Our results suggest that it might be particularly useful in patients with AIS without hypertension, although this requires confirmation in independent datasets.

## Acknowledgments

We want to acknowledge all the participants and investigators of BioBank Japan. Supercomputing resources were provided by the Human Genome Center, Institute of Medical Science, and the University of Tokyo (http://sc.hgc.jp/shirokane.html). The ToMMo summary statistics were derived from jMorp (https://jmorp.megabank.tohoku.ac.jp/gwas-studies/TGA000007). The MEGASTROKE project received funding from sources specified at https://www.megastroke.org/acknowledgements.html

## Data availability

The weights of metaGRS derived in this study will be publicly available after acceptance. Genotype datasets were deposited in the National Bioscience Database Center Human Database (BBJ1, Research ID: hum0014; BBJ2, Research ID: hum0311).

## Disclosures

Y.K. holds stock of StaGen Co, Ltd.

**Supplementary Material**

Supplementary Methods

Supplementary Notes

Tables S1–S15

Figures S1–S5

## Non-standard abbreviations and acronyms

PRS: polygenic risk score
P+T: pruning and thresholding
BBJ: BioBank Japan
BBJ1: BBJ 1^st^ cohort
BBJ2: BBJ 2^nd^ cohort
ToMMo: Tohoku Medical Megabank
AIS: all ischemic stroke
IPW: inverse probability weighting
LD: linkage disequilibrium
GWAS: genome-wide association study
PC: principal component
IPW: inverse probability weight
AUC: area under the curve
LAS: large artery stroke
SVS: small vessel stroke
CES: cardioembolic stroke
TIS: transient ischemic attack
HWE: Hardy-Weinberg equilibrium
WGS: whole genome sequencing
MI: myocardial infarction
SAP: stable angina pectoris
AP: unstable angina pectoris
AF: atrial fibrillation
DM: diabetes mellitus
SM: smoking
BMI: body mass index
HE: height
SBP: systolic blood pressure
DBP: diastolic blood pressure
TC: total cholesterol
TG: triglyceride
HDL: high-density lipoprotein
LDL: low-density lipoprotein

## SUPPLEMENTARY MATERIALS

### Supplementary Methods

#### Samples

BioBank Japan (BBJ) collected DNA, serum, and medical records (clinical information) with consent from patients. The BBJ 1^st^ cohort (BBJ1) dataset contained all ischemic stroke (AIS, n=17,621) cases, including large artery stroke (LAS, n=981), small vessel stroke (SVS, n=3,108), and cardioembolic stroke (CES, n=608) cases. The patients without AIS were included as controls (n=162,317).

Testing involved the use of data from part of the BBJ 2^nd^ cohort (BBJ2) dataset, which contains information about AIS cases (n=1,470), including LAS (n=268), SVS (n=508), CES (n=122), and transient ischemic attack (TIA, n=105). All patients without AIS were included as controls (n=40,459). Among these cases, recurrent ischemic stroke (n=187) was used as a case of recurrent AIS, which included LAS (n=40), SVS (n=57), CES (n=11), and transient ischemic attack (TIA, n=20). Samples with information on the first onset date and follow-up of under 30 days were excluded; AIS cases with recurrence (n=174) were set as the case group, and AIS cases without recurrence (n=1,153) remained in the control group in the testing sample.

#### Quality control and imputation process of BBJ1 data

We removed variants with call rates <0.99, samples with call rates <0.98, non-East Asian samples, and sex-discordant samples. We used 939 samples whose genotypes were analyzed using whole-genome sequencing (WGS); we added an additional quality control based on the concordance rate between the genotyping array and WGS. We excluded variants with concordance rates <99.5% or non-reference discordance rates ≥0.5% and Hardy-Weinberg equilibrium (HWE) (p<1e-6). The 10 principal components (PCs) were calculated by applying additional quality control (QC) to the BBJ1 genotyped data. We removed samples from close relatives (King’s cutoff >0.0884), and 24 long LD regions,^54^ including the MHC region (chr6, position 25,000,000-35,000,000), and MAF<0.01. We pruned (PLINK2 parameters: --indep-pairwise 200 50 0.20) and finally used 92,231–92,303 variants to conduct projection and calculate the first 10 PCs depending on a 10-fold sample set of the 90% BBJ1 dataset.

Subsequently, the datasets were phased (Eagle v2.4.1) and imputed (Minimac4 v1.0.2) using the developed panel.^55^ We further conducted quality control to remove variants with minor allele counts <10, close relatives (King cutoff >0.0884), and imputation r-square <0.8.

#### Quality control and imputation process for BBJ2 data

We removed samples with no age/sex information, sex discrepancy, call rates <0.98, heterozygosity rates with SD >4 or <-4, from duplicate or twins (pi-hat >= 0.75), and from non-East Asian subjects. We then removed variants with a call rate <0.99, duplicate SNPs, heterozygous count <5, HWE (p<1e-6), and allele frequency discrepancies (gap from 1000 genomes EAS >0.16). A total of 41,929 samples and 525,239 variants were analyzed.

We applied additional quality control to the BBJ2 genotyped data to calculate the 10 PCs. We removed variants in 24 long LD regions,^54^ pruned them (PLINK2 parameters: --indep-pairwise 200 50 0.05), extracted close relatives (King cut-off >0.0884), and used 69,068 variants to calculate the first 10 PCs.

We removed variants of the imputation r-squared <0.3 after phasing (Eagle v2.4.1) and imputing (Minimac4 v1.0.2) the developed panel.^55^ We used a lower r-square threshold for BBJ2 to reduce the number of variants unmatched with BBJ1.

#### Polygenic risk score parameters at the derivation

Pruning and threshold used a total of 1,224 parameter combinations of three stricter imputation r-squared score thresholds {0.8, 0.9, and 0.95}, four base sizes of the clumping window {50, 100, 200, and 500}, six squared correlations of clumping {0.01, 0.05, 0.2, 0.5, 0.8, and 0.95}, and 17 p-value threshold {1e-8, 3e-8, 1e-7, 3e-7, 1e-6, 3e-6, 1e-5, 3e-5, 1e-4, 3e-4, 1e-3, 3e-3, 0.01, 0.03, 0.1, 0.3, and 1}. We used clumping windows and divided the base size by the squared correlation of clumping.^56–59^ The following default parameters were used for LDpred2: three heritability {0.7, 1, and 1.4}×h^2^LDSC, where h^2^LDSC is the heritability estimate from the constrained LD score regression,^52^ 21 ρ estimates {equally spaced on a log scale between 1e-5 and 1}, and sparse or not. The following default parameters were used for Lassosum2: 10 values of s {0.1, 0.2, 0.3, 0.4, 0.5, 0.6, 0.7, 0.8, 0.9, and 1.0} and 20 values of λ {equally spaced on a log scale between 0.1 and 0.001}. Regarding PRS-CS and PRS-CSx, we used slightly more extended parameters: phi {1e-7, 1e-6, 1e-5, 1e-4, 1e-3, 0.01, 0.1, 1, and auto}, than the default parameters {1e-4, 1e-3, 0.01, 0.1, 1, and auto}, because the optimized parameter in the initial trials using our dataset was the smallest (phi=1e-4) in the default range. Other parameters were set to default values (a=1, b=0.5).^41,42^

#### Linkage disequilibrium (LD) reference

We used the EAS superpopulation of the 1000 Genomes panel (n=504) as a LD reference for P+T clumping. For Ldpred2, Lassosum2, PRS-CS, and PRS-CSx models, we restricted our use of external LD reference panels to the HapMap 3 variants, which were also constructed from 1000 Genomes EAS. HapMap 3 variant restriction resulted in 898,456, 898,456, 985,440, and 1,076,835 variants of Ldpred2, Lassosum2, PRS-CS, and PRS-CSx, respectively. To use PRS-CSx for the EUR, we used the EUR superpopulation of the 1000 Genomes panel and its HapMap3 variants (1,016,745 variants).

#### Genome wide association study summary statistics for metaGRS construction

MetaGRS was developed following a previous study^14^ and included nine binary and eight quantitative traits. The nine binary traits were SVS, LAS, CES, myocardial infarction (MI), stable angina pectoris (SAP), unstable angina pectoris (AP), atrial fibrillation (AF), diabetes (DM), and ever smoking (SM) from the BBJ1 dataset. Subsequently, we conducted GWAS. The number of cases and controls is listed in Table S15. Summary statistics were obtained from the jMorp website (https://jmorp.megabank.tohoku.ac.jp) and were used for eight quantitative traits—body mass index (BMI), height (HE), systolic blood pressure (SBP), diastolic blood pressure (DBP), total cholesterol (TC), triglyceride (TG), high-density lipoprotein (HDL), and low-density lipoprotein (LDL) from the Tohoku Medical Megabank Project (ToMMo; 22,033 to 47,056 samples).^60^

#### Polygenic risk score calculation from other milestone studies

We obtained effect weights from the PGS catalog (PGS000665 and PGS002725 of https://www.pgscatalog.org for PRS32 and iPGSEAS, respectively) and for PRS89 from the supplementary tables.^18^ We used variants that matched with the BBJ2 dataset (imputation r-squared > 0.3). The unmatched variants in PRS32 and PRS89 included proxy variants that showed the highest r-squared values with the index variants only from an r-squared value greater than 0.3. The r-squared values were calculated using the plink --r2 command^35^ with the 1000 Genome EAS as a reference.

### Supplementary Notes

#### Characteristics of the PRS methods during validation

The following characteristics of each method were observed while testing several parameters: the best mean incremental R^2^ of 0.0038 was observed for the P + T method when we liberalized the p-value threshold to 1 and clumped r^2^ >0.8, whereas it was below 0.001 when we used a low p-value threshold (<10^-8^, Table S5). For LDpred2, *ρ* values higher than 0.001 showed good performance (mean incremental R^2^ was 0.0038 for *ρ* values >=0.001 compared to 0.0016 for *ρ* values <0.001), while heritability estimates and sparse parameters made less difference (Table S6). For Lassosum2, the closer the value of parameter “s” is to 1, the higher the prediction accuracy, but it wasn’t the case at exactly 1. Larger lambda parameters correlate with greater accuracy (mean incremental R^2^ was between 0.0025 to 0.0039). However, the accuracy sharply decreases (mean incremental R^2^<0.001) if the value is too small (<0.01). The prediction performance was maximized when the parameters were s=0.9 and lambda=0.0038 (Table S7). High and low phi values resulted in low performance for PRS-CS (mean incremental R^2^ was 0.0041 for phi=10^-3^, 10^-4^, and 10^-5^ compared to 0.0029 for other phi parameters; Table S8). Meanwhile, PRS-CSx was relatively consistent regardless of the phi values (mean incremental R^2^ was 0.0032 for phi=10^-3^, 10^-4^, and 10^-5^ compared to 0.0028 for other phi parameters; Table S9).

The performance of the PRS-CS (R^2^=0.00441) was comparable to that of LDpred2 (R^2^=0.00443) in the validation analysis. Low validation predictability (incremental R^2^ < 0.001) was observed when we restricted the p-value threshold in the P+T method to genome-wide significance (p<5×10^-8^). PRS-CSx improves cross-population polygenic prediction by integrating GWAS summary statistics from other populations;^61–65^ however, we could not reproduce this result in our current study using PRS-CSx. Our results demonstrate the importance of using Bayesian methods of high-dimensional techniques in variable selection and shrinkage estimation considering LD (such as LDpred2 and PRS-CS) to predict AIS and recurrent AIS.

### Supplementary Figures

**Figure S1.**
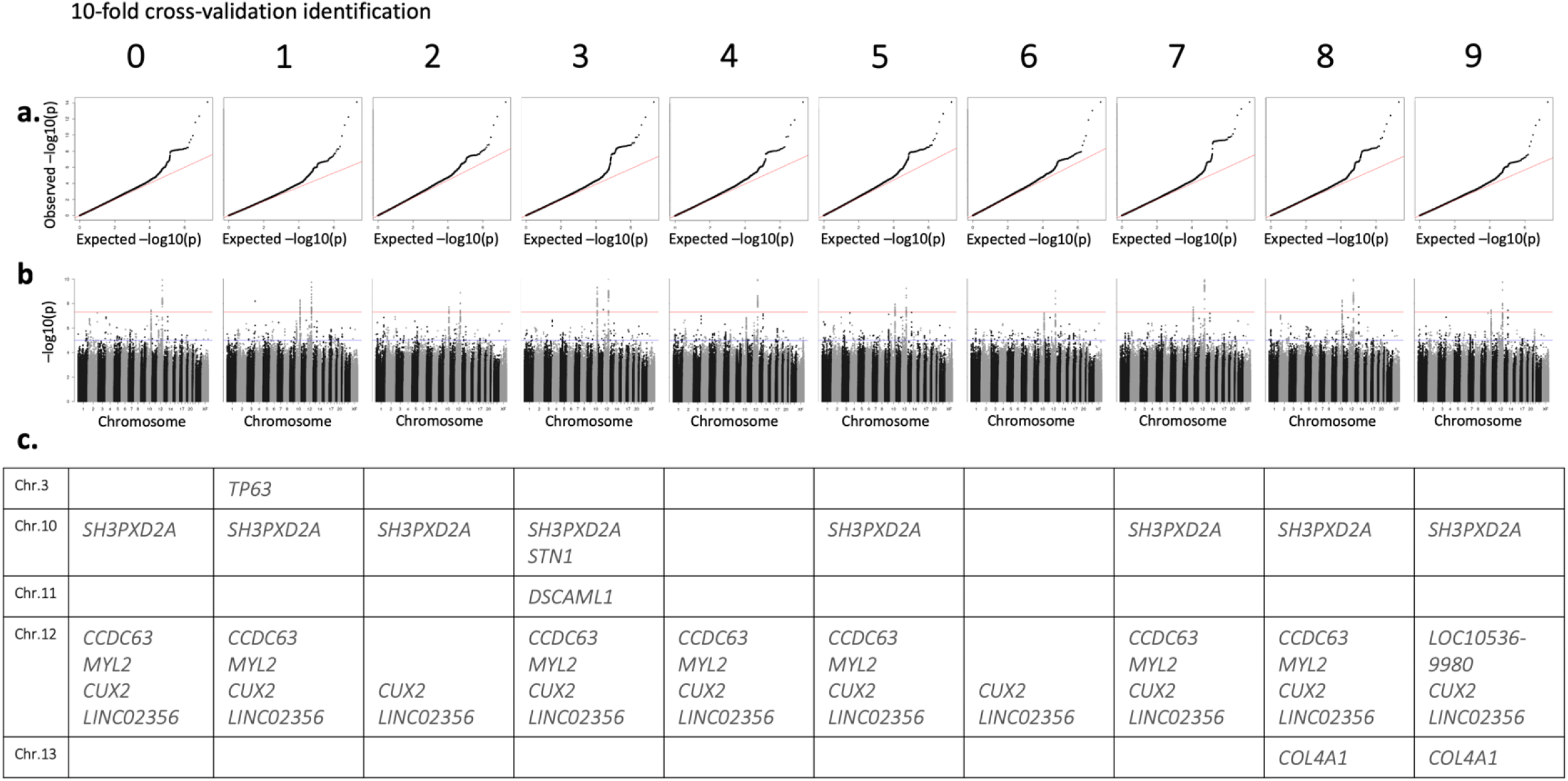
Genome-wide association study (GWAS) in derivation samples. We conducted stroke GWAS using 90% of BBJ 1st cohort data as the derivation sample, using Firth logistic regression and PLINK (v.2.0) for each 10-fold cross-validation identification. a. Quantile-Quantile plot, b. Manhattan plot, and c. genome-wide significant (p<5e-8) loci of each fold. Abbreviations: Chr = chromosome.

**Figure S2.**
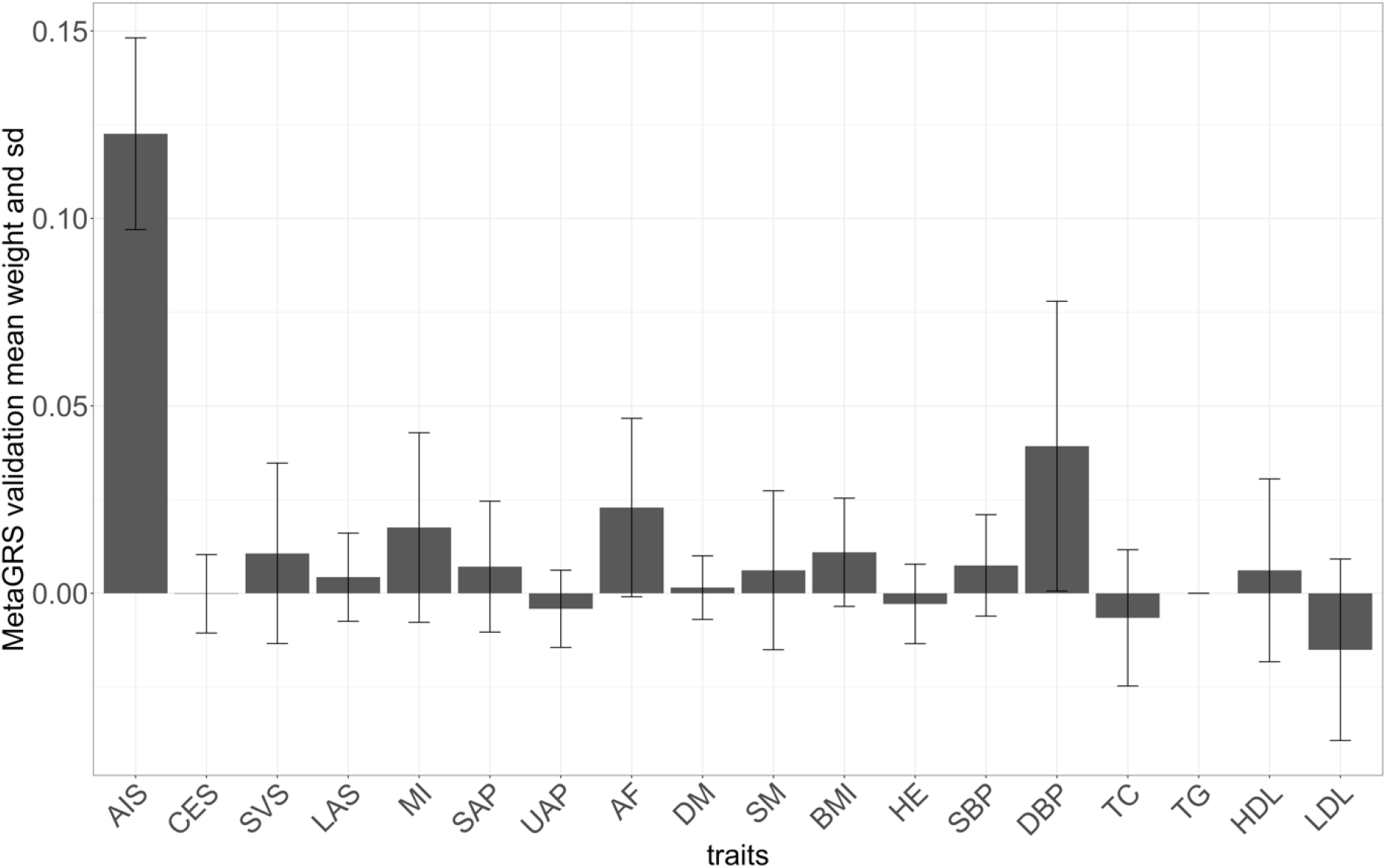
Elastic net weight. The mean weight of 10-fold elastic net regression as determined by “glmnet” in the validation sample. The X-axis shows the AS and the 17 binary and quantitative traits. Error bars represent standard deviations. Abbreviations: AIS, all ischemic stroke; SVS, small vessel stroke; LAS, large artery stroke; CES, cardioembolic stroke; MI, myocardial infarction; SAP, stable angina pectoris; UAP, unstable angina pectoris; AF, atrial fibrillation; DM, diabetes; SM, smoking; BMI, body mass index; HE, height; SBP, systolic blood pressure; DBP, diastolic blood pressure; TC, total cholesterol; TG, triglyceride; HCL, high-density lipoprotein; LDL, low-density lipoprotein; SD, standard deviation.

**Figure S3.**
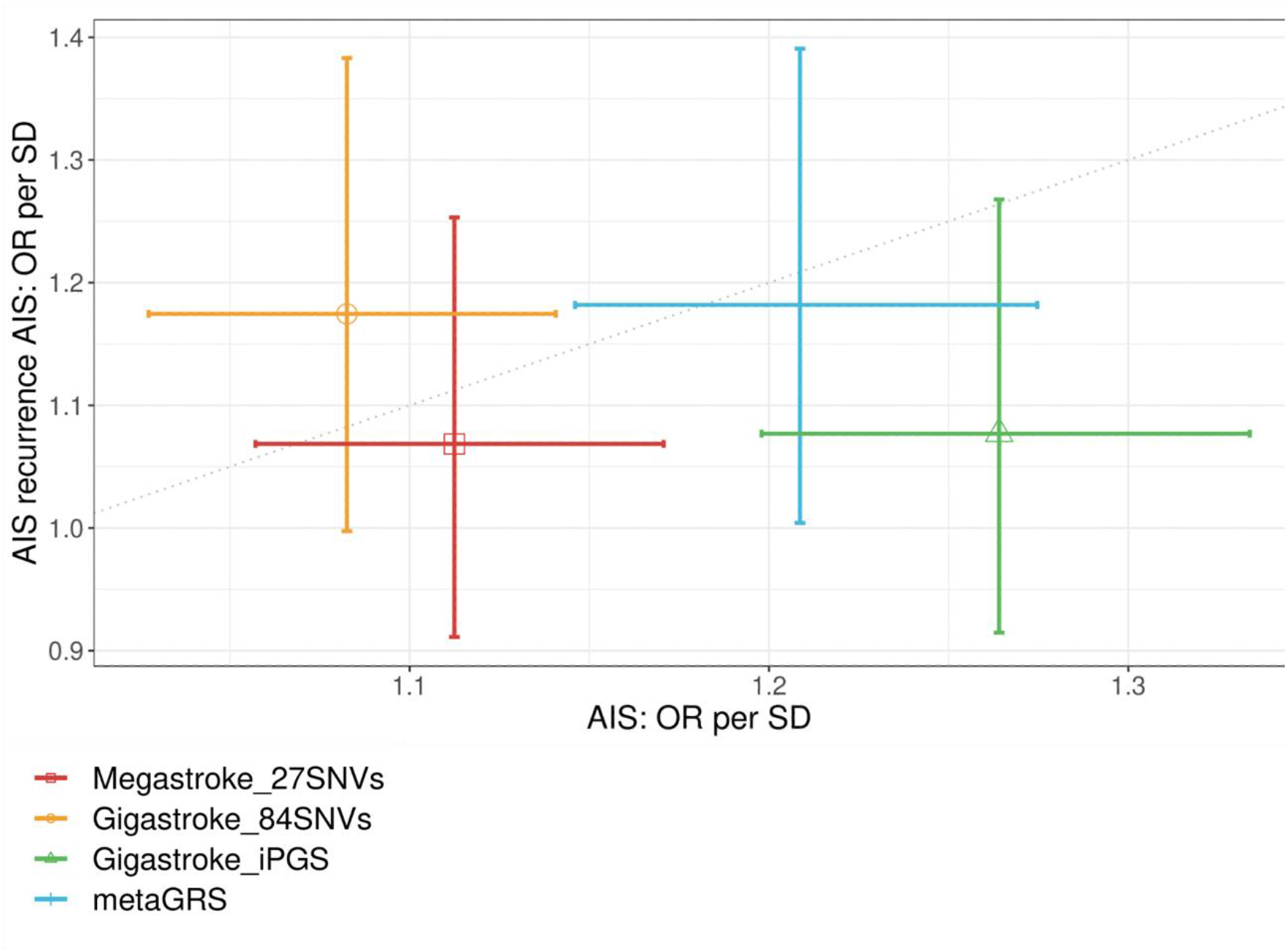
Odds ratio per SD to predict AIS and AIS recurrence. Odds ratio per standard deviation by metaGRS and three publicly available PRS in the independent test set of AIS (case=1, 470, control=40,459) and AIS recurrence case-control set (case=174, control=1,153). Age, sex, the first 10 PCs, and seven risk factors were adjusted. Error bars represent 95% confidence intervals. The dotted line represents the point at which the ORs per SD of AIS and AIS recurrence were equal. Abbreviations: OR, odds ratio; AIS, all ischemic stroke; PRS, polygenic risk score; PC, principal component; SD, standard deviation.

**Figure S4.**
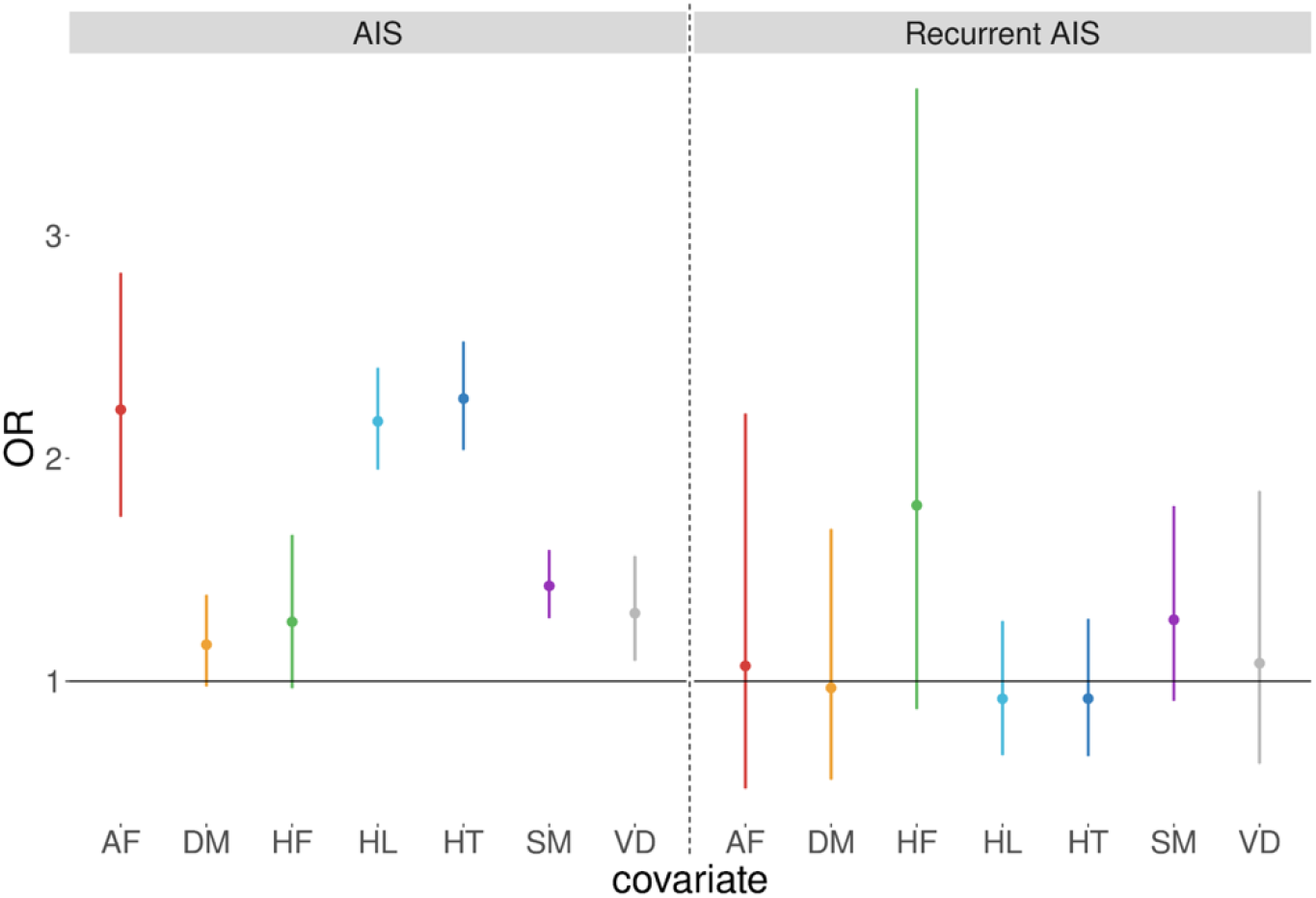
Association between seven comorbidities and AIS/AIS recurrence. The Y-axis shows the odds ratio per standard deviation (SD) and 95% confidence interval (CI) of the seven risk factors (six clinical comorbidities and smoking) in the BBJ 2^nd^ cohort. In our dataset, we used HT defined as SBP>140 mmHg, DBP>90 mmHg, or hypertension history, DM inclusive of type 1 diabetes and other diabetes, SM as a current smoker at the time of registration, VD representing myocardial infarction, arteriosclerosis obliterans, stable angina pectoris, unstable angina pectoris, and AF inclusive of atrial flutter. We used the status at the time of registration and history of these comorbidities. Abbreviations: HT, hypertension; HL, hyperlipidemia; DM, diabetes; SM, smoking; VD, vascular disease; HF, congestive heart failure; AF, atrial fibrillation.

**Figure S5.**
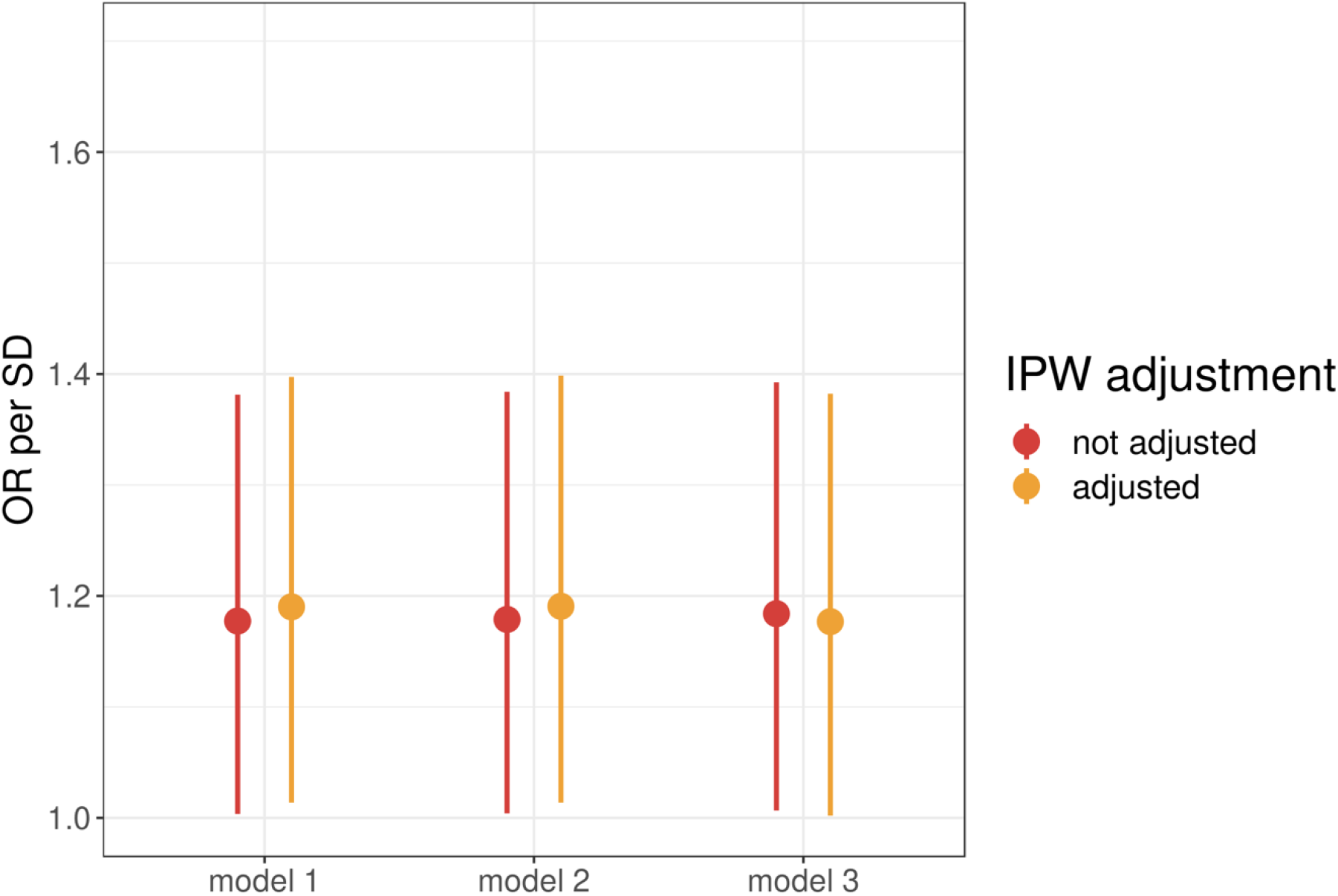
Association between metaGRS and AIS recurrence in patients with or without IPW adjustment. The y-axis shows the odds ratio per SD and the 95% confidence interval for predicting recurrent AIS. Model 1: age and sex; model 2: age, sex, and seven risk factors; model 3: age, sex, the first 10 principal components (PCs), and seven risk factors. The color represents whether the inverse probability weight (IPW) is adjusted. We applied logistic regression using variables as covariates when the IPW was not adjusted. We replaced 14 missing data on the smoking status to mean values.

### Supplementary Tables

In separate spreadsheets S1-15

## Consortium authors

### BioBank Japan

Koichi Matsuda^1,2^, Yuji Yamanashi^3^, Yoichi Furukawa^4^, Takayuki Morisaki^5^, Yoshinori Murakami^6^, Yoichiro Kamatani^2,7^, Kaori Muto^8^, Akiko Nagai^8^, Wataru Obara^9^, Ken Yamaji^10^, Kazuhisa Takahashi^11^, Satoshi Asai^12,13^, Yasuo Takahashi^13^, Takao Suzuki^14^, Nobuaki Sinozaki^14^, Hiroki Yamaguchi^15^, Shiro Minami^16^, Shigeo Murayama^17^, Kozo Yoshimori^18^, Satoshi Nagayama^19^, Daisuke Obata^20^, Masahiko Higashiyama^21^, Akihide Masumoto^22^, Yukihiro Koretsune^23^

^1^Laboratory of Genome Technology, Human Genome Center, Institute of Medical Science, The University of Tokyo, Tokyo, Japan.

^2^Laboratory of Clinical Genome Sequencing, Graduate School of Frontier Sciences, The

University of Tokyo, Tokyo, Japan.

^3^Division of Genetics, The Institute of Medical Science, The University of Tokyo, Tokyo, Japan.

^4^Division of Clinical Genome Research, Institute of Medical Science, The University of

Tokyo, Tokyo, Japan.

^5^Division of Molecular Pathology IMSUT Hospital, Department of Internal Medicine Project Division of Genomic Medicine and Disease Prevention The Institute of Medical Science The University of Tokyo, Tokyo, Japan.

^6^Department of Cancer Biology, Institute of Medical Science, The University of Tokyo,

Tokyo, Japan.

^7^Laboratory of Complex Trait Genomics, Graduate School of Frontier Sciences, The University of Tokyo, Tokyo, Japan.

^8^Department of Public Policy, Institute of Medical Science, The University of Tokyo, Tokyo, Japan.

^9^Department of Urology, Iwate Medical University, Iwate, Japan.

^10^Department of Internal Medicine and Rheumatology, Juntendo University Graduate School of Medicine, Tokyo, Japan.

^11^Department of Respiratory Medicine, Juntendo University Graduate School of Medicine,

Tokyo, Japan.

^12^Division of Pharmacology, Department of Biomedical Science, Nihon University School of Medicine, Tokyo, Japan.

^13^Division of Genomic Epidemiology and Clinical Trials, Clinical Trials Research Center,

Nihon University. School of Medicine, Tokyo, Japan.

^14^Tokushukai Group, Tokyo, Japan.

^15^Hiroki Yamaguchi. Department of Hematology, Nippon Medical School, Tokyo, Japan.

^16^Department of Bioregulation, Nippon Medical School, Kawasaki, Japan.

^17^Shigeo Murayama. Tokyo Metropolitan Geriatric Hospital and Institute of Gerontology, Tokyo, Japan.

^18^Fukujuji Hospital, Japan Anti-Tuberculosis Association, Tokyo, Japan.

^19^The Cancer Institute Hospital of the Japanese Foundation for Cancer Research, Tokyo, Japan.

^20^Center for Clinical Research and Advanced Medicine, Shiga University of Medical Science, Shiga, Japan.

^21^Masahiko Higashiyama. Department of General Thoracic Surgery, Osaka International Cancer Institute, Osaka, Japan.

^22^Iizuka Hospital, Fukuoka, Japan.

^23^National Hospital Organization Osaka National Hospital, Osaka, Japan.

### MEGASTROKE CONSORTIUM

Rainer Malik^1^, Ganesh Chauhan^2^, Matthew Traylor^3^, Muralidharan Sargurupremraj^4,5^, Yukinori Okada^6,7,8^, Aniket Mishra^4,5^, Loes Rutten-Jacobs^3^, Anne-Katrin Giese^9^, Sander W van der Laan^10^, Solveig Gretarsdottir^11^, Christopher D Anderson^12,13,14^, Michael Chong^15^, Hieab HH Adams^16,17^, Tetsuro Ago^18^, Peter Almgren^19^, Philippe Amouyel^20,21^, Hakan Ay^13,22^, Traci M Bartz^23^, Oscar R Benavente^24^, Steve Bevan^25^, Giorgio B Boncoraglio^26^, Robert D Brown, Jr.^27^, Adam S Butterworth^28,29^, Caty Carrera^30,31^, Cara L Carty^32,33^, Daniel I Chasman^34,35^, Wei-Min Chen^36^, John W Cole^37^, Adolfo Correa^38^, Ioana Cotlarciuc^39^, Carlos Cruchaga^40,41^, John Danesh^28,42,43,44^ , Paul IW de Bakker^45,46^, Anita L DeStefano^47,48^, Marcel den Hoed^49^, Qing Duan^50^, Stefan T Engelter^51,52^, Guido J Falcone^53,54^, Rebecca F Gottesman^55^, Raji P Grewal^56^, Vilmundur Gudnason^57,58^, Stefan Gustafsson^59^, Jeffrey Haessler^60^, Tamara B Harris^61^, Ahamad Hassan^62^, Aki S Havulinna^63,64^, Susan R Heckbert^65^, Elizabeth G Holliday^66,67^, George Howard^68^, Fang-Chi Hsu^69^, Hyacinth I Hyacinth^70^, M Arfan Ikram^16^, Erik Ingelsson^71,72^, Marguerite R Irvin^73^, Xueqiu Jian^74^, Jordi Jiménez-Conde^75^, Julie A Johnson^76,77^, J Wouter Jukema^78^, Masahiro Kanai^6,7,79^, Keith L Keene^80,81^, Brett M Kissela^82^, Dawn O Kleindorfer^82^, Charles Kooperberg^60^, Michiaki Kubo^83^, Leslie A Lange^84^, Carl D Langefeld^85^, Claudia Langenberg^86^, Lenore J Launer^87^, Jin-Moo Lee^88^, Robin Lemmens^89,90^, Didier Leys^91^, Cathryn M Lewis^92,93^, Wei-Yu Lin^28,94^, Arne G Lindgren^95,96^, Erik Lorentzen^97^, Patrik K Magnusson^98^, Jane Maguire^99^, Ani Manichaikul^36^, Patrick F McArdle^100^, James F Meschia^101^, Braxton D Mitchell^100,102^, Thomas H Mosley^103,104^, Michael A Nalls^105,106^, Toshiharu Ninomiya^107^, Martin J O’Donnell^15,108^, Bruce M Psaty^109,110,111,112^ , Sara L Pulit^45,113^, Kristiina Rannikmäe^114,115^, Alexander P Reiner^65,116^, Kathryn M Rexrode^117^, Kenneth Rice^118^, Stephen S Rich^36^, Paul M Ridker^34,35^, Natalia S Rost^9,13^, Peter M Rothwell^119^, Jerome I Rotter^120,121^, Tatjana Rundek^122^, Ralph L Sacco^122^, Saori Sakaue^7,123^, Michele M Sale^124^, Veikko Salomaa^63^, Bishwa R Sapkota^125^, Reinhold Schmidt^126^, Carsten O Schmidt^127^, Ulf Schminke^128^, Pankaj Sharma^39^, Agnieszka Slowik^129^, Cathie LM Sudlow^114,115^, Christian Tanislav^130^, Turgut Tatlisumak^131,132^, Kent D Taylor^120,121^, Vincent NS Thijs^133,134^, Gudmar Thorleifsson^11^, Unnur Thorsteinsdottir^11^, Steffen Tiedt^1^, Stella Trompet^135^, Christophe Tzourio^5,136,137^, Cornelia M van Duijn^138,139^, Matthew Walters^140^, Nicholas J Wareham^86^, Sylvia Wassertheil-Smoller^141^, James G Wilson^142^, Kerri L Wiggins^109^, Qiong Yang^47^, Salim Yusuf^15^, Najaf Amin^16^, Hugo S Aparicio^48,185^, Donna K Arnett^186^, John Attia^187^, Alexa S Beiser^47,48^, Claudine Berr^188^, Julie E Buring^34,35^, Mariana Bustamante^189^, Valeria Caso^190^, Yu-Ching Cheng^191^, Seung Hoan Choi^48,192^, Ayesha Chowhan^48,185^, Natalia Cullell^31^, Jean-François Dartigues^193,194^, Hossein Delavaran^95,96^, Pilar Delgado^195^, Marcus Dörr^196,197^, Gunnar Engström^19^, Ian Ford^198^, Wander S Gurpreet^199^, Anders Hamsten ^200,201^, Laura Heitsch^202^, Atsushi Hozawa^203^, Laura Ibanez^204^, Andreea Ilinca^95,96^, Martin Ingelsson^205^, Motoki Iwasaki^206^, Rebecca D Jackson^207^, Katarina Jood^208^, Pekka Jousilahti^63^, Sara Kaffashian^4,5^, Lalit Kalra^209^, Masahiro Kamouchi^210^, Takanari Kitazono^211^, Olafur Kjartansson^212^, Manja Kloss^213^, Peter J Koudstaal^214^, Jerzy Krupinski^215^, Daniel L Labovitz^216^, Cathy C Laurie^118^, Christopher R Levi^217^, Linxin Li^218^, Lars Lind^219^, Cecilia M Lindgren^220,221^, Vasileios Lioutas^48,222^, Yong Mei Liu^223^, Oscar L Lopez^224^, Hirata Makoto^225^, Nicolas Martinez-Majander^172^, Koichi Matsuda^225^, Naoko Minegishi^203^, Joan Montaner^226^, Andrew P Morris^227,228^, Elena Muiño^31^, Martina Müller-Nurasyid^229,230,231^ , Bo Norrving^95,96^, Soichi Ogishima^203^, Eugenio A Parati^232^, Leema Reddy Peddareddygari^56^, Nancy L Pedersen^98,233^, Joanna Pera^129^, Markus Perola^63,234^, Alessandro Pezzini^235^, Silvana Pileggi^236^, Raquel Rabionet^237^, Iolanda Riba-Llena^30^, Marta Ribasés^238^, Jose R Romero^48,185^, Jaume Roquer^239,240^, Anthony G Rudd^241,242^, Antti-Pekka Sarin^243,244^, Ralhan Sarju^199^, Chloe Sarnowski^47,48^, Makoto Sasaki^245^, Claudia L Satizabal^48,185^, Mamoru Satoh^245^, Naveed Sattar^246^, Norie Sawada^206^, Gerli Sibolt^172^, Ásgeir Sigurdsson^247^, Albert Smith^248^, Kenji Sobue^245^, Carolina Soriano-Tárraga^240^, Tara Stanne^249^, O Colin Stine^250^, David J Stott^251^, Konstantin Strauch^229,252^, Takako Takai^203^, Hideo Tanaka^253,254^, Kozo Tanno^245^, Alexander Teumer^255^, Liisa Tomppo^172^, Nuria P Torres-Aguila^31^, Emmanuel Touze^256,257^, Shoichiro Tsugane^206^, Andre G Uitterlinden^258^, Einar M Valdimarsson^259^, Sven J van der Lee^16^, Henry Völzke^255^, Kenji Wakai^253^, David Weir^260^, Stephen R Williams^261^, Charles DA Wolfe^241,242^, Quenna Wong^118^, Huichun Xu^191^, Taiki Yamaji^206^, Dharambir K Sanghera^125,169,170^ , Olle Melander^19^, Christina Jern^171^, Daniel Strbian^172,173^, Israel Fernandez-Cadenas^31,30^, W T Longstreth, Jr^65,174^, Arndt Rolfs^175^, Jun Hata^107^, Daniel Woo^82^, Jonathan Rosand^12,13,14^, Guillaume Pare^15^, Jemma C Hopewell^176^, Danish Saleheen^177^, Kari Stefansson^11,178^, Bradford B Worrall^179^, Steven J Kittner^37^, Sudha Seshadri^48,180^, Myriam Fornage^74,181^, Hugh S Markus^3^, Joanna MM Howson^28^, Yoichiro Kamatani^6,182^, Stephanie Debette^4,5^, Martin Dichgans^1,183,184^

^1^Institute for Stroke and Dementia Research (ISD), University Hospital, LMU Munich, Munich, Germany

^2^Centre for Brain Research, Indian Institute of Science, Bangalore, India

^3^Stroke Research Group, Division of Clinical Neurosciences, University of Cambridge, UK

^4^INSERM U1219 Bordeaux Population Health Research Center, Bordeaux, France

^5^University of Bordeaux, Bordeaux, France

^6^Laboratory for Statistical Analysis, RIKEN Center for Integrative Medical Sciences, Yokohama, Japan

^7^Department of Statistical Genetics, Osaka University Graduate School of Medicine, Osaka, Japan

^8^Laboratory of Statistical Immunology, Immunology Frontier Research Center (WPI-IFReC), Osaka University, Suita, Japan.

^9^Department of Neurology, Massachusetts General Hospital, Harvard Medical School, Boston, MA, USA

^10^Laboratory of Experimental Cardiology, Division of Heart and Lungs, University Medical Center Utrecht, University of Utrecht, Utrecht,Netherlands

^11^deCODE genetics/AMGEN inc, Reykjavik, Iceland

^12^Center for Genomic Medicine, Massachusetts General Hospital (MGH), Boston, MA, USA

^13^J. Philip Kistler Stroke Research Center, Department of Neurology, MGH, Boston, MA, USA

^14^Program in Medical and Population Genetics, Broad Institute, Cambridge, MA, USA

^15^Population Health Research Institute, McMaster University, Hamilton, Canada

^16^Department of Epidemiology, Erasmus University Medical Center, Rotterdam, Netherlands

^17^Department of Radiology and Nuclear Medicine, Erasmus University Medical Center, Rotterdam, Netherlands

^18^Department of Medicine and Clinical Science, Graduate School of Medical Sciences, Kyushu University, Fukuoka, Japan

^19^Department of Clinical Sciences, Lund University, Malmö, Sweden

^20^Univ. Lille, Inserm, Institut Pasteur de Lille, LabEx DISTALZ-UMR1167, Risk factors and molecular determinants of aging-related diseases, F-59000 Lille, France

^21^Centre Hosp. Univ Lille, Epidemiology and Public Health Department, F-59000 Lille, France

^22^AA Martinos Center for Biomedical Imaging, Department of Radiology, Massachusetts General Hospital, Harvard Medical School, Boston, MA, USA

^23^Cardiovascular Health Research Unit, Departments of Biostatistics and Medicine, University of Washington, Seattle, WA, USA

^24^Division of Neurology, Faculty of Medicine, Brain Research Center, University of British Columbia, Vancouver, Canada

^25^School of Life Science, University of Lincoln, Lincoln, UK

^26^Department of Cerebrovascular Diseases, Fondazione IRCCS Istituto Neurologico "Carlo Besta", Milano, Italy

^27^Department of Neurology, Mayo Clinic Rochester, Rochester, MN, USA

^28^MRC/BHF Cardiovascular Epidemiology Unit, Department of Public Health and Primary Care, University of Cambridge, Cambridge, UK

^29^The National Institute for Health Research Blood and Transplant Research Unit in Donor Health and Genomics, University of Cambridge, UK

^30^Neurovascular Research Laboratory, Vall d’Hebron Institut of Research, Neurology and Medicine Departments-Universitat Autònoma de Barcelona, Vall d’Hebrón Hospital, Barcelona, Spain

^31^Stroke Pharmacogenomics and Genetics, Fundacio Docència i Recerca MutuaTerrassa, Terrassa, Spain

^32^Children’s Research Institute, Children’ s National Medical Center, Washington, DC, USA

^33^Center for Translational Science, George Washington University, Washington, DC, USA

^34^Division of Preventive Medicine, Brigham and Women’s Hospital, Boston, MA, USA

^35^Harvard Medical School, Boston, MA, USA

^36^Center for Public Health Genomics, Department of Public Health Sciences, University of Virginia, Charlottesville, VA, USA

^37^Department of Neurology, University of Maryland School of Medicine and Baltimore VAMC, Baltimore, MD, USA

^38^Departments of Medicine, Pediatrics and Population Health Science, University of Mississippi Medical Center, Jackson, MS, USA

^39^Institute of Cardiovascular Research, Royal Holloway University of London, UK & Ashford and St Peters Hospital, Surrey UK

^40^Department of Psychiatry,The Hope Center Program on Protein Aggregation and Neurodegeneration (HPAN),Washington University, School of Medicine, St. Louis, MO, USA

^41^Department of Developmental Biology, Washington University School of Medicine, St. Louis, MO, USA

^42^NIHR Blood and Transplant Research Unit in Donor Health and Genomics, Department of Public Health and Primary Care, University of Cambridge, Cambridge, UK

^43^Wellcome Trust Sanger Institute, Wellcome Trust Genome Campus, Hinxton, Cambridge, UK

^44^British Heart Foundation, Cambridge Centre of Excellence, Department of Medicine, University of Cambridge, Cambridge, UK

^45^Department of Medical Genetics, University Medical Center Utrecht, Utrecht, Netherlands

^46^Department of Epidemiology, Julius Center for Health Sciences and Primary Care, University Medical Center Utrecht, Utrecht, Netherlands

^47^Boston University School of Public Health, Boston, MA, USA

^48^Framingham Heart Study, Framingham, MA, USA

^49^Department of Immunology, Genetics and Pathology and Science for Life Laboratory, Uppsala University, Uppsala, Sweden

^50^Department of Genetics, University of North Carolina, Chapel Hill, NC, USA

^51^Department of Neurology and Stroke Center, Basel University Hospital, Switzerland

^52^Neurorehabilitation Unit, University and University Center for Medicine of Aging and Rehabilitation Basel, Felix Platter Hospital, Basel, Switzerland

^53^Department of Neurology, Yale University School of Medicine, New Haven, CT, USA

^54^Program in Medical and Population Genetics, The Broad Institute of Harvard and MIT, Cambridge, MA, USA

^55^Department of Neurology, Johns Hopkins University School of Medicine,

Baltimore, MD, USA

^56^Neuroscience Institute, SF Medical Center, Trenton, NJ, USA

^57^Icelandic Heart Association Research Institute, Kopavogur, Iceland

^58^University of Iceland, Faculty of Medicine, Reykjavik, Iceland

^59^Department of Medical Sciences, Molecular Epidemiology and Science for Life Laboratory, Uppsala University, Uppsala, Sweden

^60^Division of Public Health Sciences, Fred Hutchinson Cancer Research Center, Seattle, WA, USA

^61^Laboratory of Epidemiology and Population Science, National Institute on Aging, National Institutes of Health, Bethesda, MD, USA

^62^Department of Neurology, Leeds General Infirmary, Leeds Teaching Hospitals NHS Trust, Leeds, UK

^63^National Institute for Health and Welfare, Helsinki, Finland

^64^FIMM - Institute for Molecular Medicine Finland, Helsinki, Finland

^65^Department of Epidemiology, University of Washington, Seattle, WA, USA

^66^Public Health Stream, Hunter Medical Research Institute, New Lambton, Australia

^67^Faculty of Health and Medicine, University of Newcastle, Newcastle, Australia

^68^School of Public Health, University of Alabama at Birmingham, Birmingham, AL, USA

^69^Department of Biostatistical Sciences, Wake Forest School of Medicine, Winston-Salem, NC, USA

^70^Aflac Cancer and Blood Disorder Center, Department of Pediatrics, Emory University School of Medicine, Atlanta, GA, USA

^71^Department of Medicine, Division of Cardiovascular Medicine, Stanford University School of Medicine, CA, USA

^72^Department of Medical Sciences, Molecular Epidemiology and Science for Life Laboratory, Uppsala University, Uppsala, Sweden

^73^Epidemiology, School of Public Health, University of Alabama at Birmingham, USA

^74^Brown Foundation Institute of Molecular Medicine, University of Texas Health Science Center at Houston, Houston, TX, USA

^75^Neurovascular Research Group (NEUVAS), Neurology Department, Institut Hospital del Mar d’Investigació Mèdica, Universitat Autònoma de Barcelona, Barcelona, Spain

^76^Department of Pharmacotherapy and Translational Research and Center for Pharmacogenomics, University of Florida, College of Pharmacy, Gainesville, FL, USA

^77^Division of Cardiovascular Medicine, College of Medicine, University of Florida, Gainesville, FL, USA

^78^Department of Cardiology, Leiden University Medical Center, Leiden, the Netherlands

^79^Program in Bioinformatics and Integrative Genomics, Harvard Medical School, Boston, MA, USA

^80^Department of Biology, East Carolina University, Greenville, NC, USA

^81^Center for Health Disparities, East Carolina University, Greenville, NC, USA

^82^University of Cincinnati College of Medicine, Cincinnati, OH, USA

^83^RIKEN Center for Integrative Medical Sciences, Yokohama, Japan

^84^Department of Medicine, University of Colorado Denver, Anschutz Medical Campus, Aurora, CO, USA

^85^Center for Public Health Genomics and Department of Biostatistical Sciences, Wake Forest School of Medicine, Winston-Salem, NC, USA

^86^MRC Epidemiology Unit, University of Cambridge School of Clinical Medicine, Institute of Metabolic Science, Cambridge Biomedical Campus, Cambridge, UK

^87^Intramural Research Program, National Institute on Aging, National Institutes of Health, Bethesda, MD, USA

^88^Department of Neurology, Radiology, and Biomedical Engineering, Washington University School of Medicine, St. Louis, MO, USA

^89^KU Leuven – University of Leuven, Department of Neurosciences, Experimental Neurology, Leuven, Belgium

^90^VIB Center for Brain & Disease Research, University Hospitals Leuven, Department of Neurology, Leuven, Belgium

^91^Univ.-Lille, INSERM U 1171. CHU Lille. Lille, France

^92^Department of Medical and Molecular Genetics, King’s College London, London, UK

^93^SGDP Centre, Institute of Psychiatry, Psychology & Neuroscience, King’s College London, London, UK

^94^Northern Institute for Cancer Research, Paul O’Gorman Building, Newcastle University, Newcastle, UK

^95^Department of Clinical Sciences Lund, Neurology, Lund University, Lund, Sweden

^96^Department of Neurology and Rehabilitation Medicine, Skåne University Hospital, Lund, Sweden

^97^Bioinformatics Core Facility, University of Gothenburg, Gothenburg, Sweden

^98^Department of Medical Epidemiology and Biostatistics, Karolinska Institutet, Stockholm, Sweden

^99^University of Technology Sydney, Faculty of Health, Ultimo, Australia

^100^Department of Medicine, University of Maryland School of Medicine, MD, USA

^101^Department of Neurology, Mayo Clinic, Jacksonville, FL, USA

^102^Geriatrics Research and Education Clinical Center, Baltimore Veterans Administration Medical Center, Baltimore, MD, USA

^103^Division of Geriatrics, School of Medicine, University of Mississippi Medical Center, Jackson, MS, USA

^104^Memory Impairment and Neurodegenerative Dementia Center, University of Mississippi Medical Center, Jackson, MS, USA

^105^Laboratory of Neurogenetics, National Institute on Aging, National institutes of Health, Bethesda, MD, USA

^106^Data Tecnica International, Glen Echo MD, USA

^107^Department of Epidemiology and Public Health, Graduate School of Medical Sciences, Kyushu University, Fukuoka, Japan

^108^Clinical Research Facility, Department of Medicine, NUI Galway, Galway, Ireland

^109^Cardiovascular Health Research Unit, Department of Medicine, University of Washington, Seattle, WA, USA

^110^Department of Epidemiology, University of Washington, Seattle, WA, USA

^111^Department of Health Services, University of Washington, Seattle, WA, USA

^112^Kaiser Permanente Washington Health Research Institute, Seattle, WA, USA

^113^Brain Center Rudolf Magnus, Department of Neurology, University Medical

Center Utrecht, Utrecht, The Netherlands

^114^Usher Institute of Population Health Sciences and Informatics, University of Edinburgh, Edinburgh, UK

^115^Centre for Clinical Brain Sciences, University of Edinburgh, Edinburgh, UK

^116^Fred Hutchinson Cancer Research Center, University of Washington, Seattle, WA, USA

^117^Department of Medicine, Brigham and Women’s Hospital, Boston, MA, USA

^118^Department of Biostatistics, University of Washington, Seattle, WA, USA

^119^Nuffield Department of Clinical Neurosciences, University of Oxford, UK

^120^Institute for Translational Genomics and Population Sciences, Los Angeles Biomedical Research Institute at Harbor-UCLA Medical Center, Torrance, CA, USA

^121^Division of Genomic Outcomes, Department of Pediatrics, Harbor-UCLA Medical Center, Torrance, CA, USA

^122^Department of Neurology, Miller School of Medicine, University of Miami, Miami, FL, USA

^123^Department of Allergy and Rheumatology, Graduate School of Medicine, the University of Tokyo, Tokyo, Japan

^124^Center for Public Health Genomics, University of Virginia, Charlottesville, VA, USA

^125^Department of Pediatrics, College of Medicine, University of Oklahoma Health Sciences Center, Oklahoma City, OK, USA

^126^Department of Neurology, Medical University of Graz, Graz, Austria

^127^University Medicine Greifswald, Institute for Community Medicine, SHIP-KEF, Greifswald, Germany

^128^University Medicine Greifswald, Department of Neurology, Greifswald, Germany

^129^Department of Neurology, Jagiellonian University, Krakow, Poland

^130^Department of Neurology, Justus Liebig University, Giessen, Germany

^131^Department of Clinical Neurosciences/Neurology, Institute of Neuroscience and Physiology, Sahlgrenska Academy at University of Gothenburg, Gothenburg, Sweden

^132^Sahlgrenska University Hospital, Gothenburg, Sweden

^133^Stroke Division, Florey Institute of Neuroscience and Mental Health, University of Melbourne, Heidelberg, Australia

^134^Austin Health, Department of Neurology, Heidelberg, Australia

^135^Department of Internal Medicine, Section Gerontology and Geriatrics, Leiden University Medical Center, Leiden, the Netherlands

^136^INSERM U1219, Bordeaux, France

^137^Department of Public Health, Bordeaux University Hospital, Bordeaux, France

^138^Genetic Epidemiology Unit, Department of Epidemiology, Erasmus University Medical Center Rotterdam, Netherlands

^139^Center for Medical Systems Biology, Leiden, Netherlands

^140^School of Medicine, Dentistry and Nursing at the University of Glasgow, Glasgow, UK

^141^Department of Epidemiology and Population Health, Albert Einstein College of Medicine, NY, USA

^142^Department of Physiology and Biophysics, University of Mississippi Medical Center, Jackson, MS, USA

^143^A full list of members and affiliations appears in the Supplementary Note

^144^Department of Human Genetics, McGill University, Montreal, Canada

^145^Department of Pathophysiology, Institute of Biomedicine and Translation Medicine, University of Tartu, Tartu, Estonia

^146^Department of Cardiac Surgery, Tartu University Hospital, Tartu, Estonia

^147^Clinical Gene Networks AB, Stockholm, Sweden

^148^Department of Genetics and Genomic Sciences, The Icahn Institute for Genomics and Multiscale Biology Icahn School of Medicine at Mount Sinai, New York, NY , USA

^149^Department of Pathophysiology, Institute of Biomedicine and Translation Medicine, University of Tartu, Biomeedikum, Tartu, Estonia

^150^Integrated Cardio Metabolic Centre, Department of Medicine, Karolinska Institutet, Karolinska Universitetssjukhuset, Huddinge, Sweden.

^151^Clinical Gene Networks AB, Stockholm, Sweden

^152^Sorbonne Universités, UPMC Univ. Paris 06, INSERM, UMR_S 1166, Team Genomics & Pathophysiology of Cardiovascular Diseases, Paris, France

^153^ICAN Institute for Cardiometabolism and Nutrition, Paris, France

^154^Department of Biomedical Engineering, University of Virginia, Charlottesville, VA, USA

^155^Group Health Research Institute, Group Health Cooperative, Seattle, WA, USA

^156^Seattle Epidemiologic Research and Information Center, VA Office of Research and Development, Seattle, WA, USA

^157^Cardiovascular Research Center, Massachusetts General Hospital, Boston, MA, USA

^158^Department of Medical Research, Bærum Hospital, Vestre Viken Hospital Trust, Gjettum, Norway

^159^Saw Swee Hock School of Public Health, National University of Singapore and National University Health System, Singapore

^160^National Heart and Lung Institute, Imperial College London, London, UK

^161^Department of Gene Diagnostics and Therapeutics, Research Institute, National Center for Global Health and Medicine, Tokyo, Japan

^162^Department of Epidemiology, Tulane University School of Public Health and Tropical Medicine, New Orleans, LA, USA

^163^Department of Cardiology, University Medical Center Groningen, University of Groningen, Netherlands

^164^MRC-PHE Centre for Environment and Health, School of Public Health, Department of Epidemiology and Biostatistics, Imperial College London, London, UK

^165^Department of Epidemiology and Biostatistics, Imperial College London, London, UK

^166^Department of Cardiology, Ealing Hospital NHS Trust, Southall, UK

^167^National Heart, Lung and Blood Research Institute, Division of Intramural Research, Population Sciences Branch, Framingham, MA, USA

^168^A full list of members and affiliations appears at the end of the manuscript

^169^Department of Pharmaceutical Sciences, College of Pharmacy, University of Oklahoma Health Sciences Center, Oklahoma City, OK, USA

^170^Oklahoma Center for Neuroscience, Oklahoma City, OK, USA

^171^Department of Pathology and Genetics, Institute of Biomedicine, The Sahlgrenska Academy at University of Gothenburg, Gothenburg, Sweden

^172^Department of Neurology, Helsinki University Hospital, Helsinki, Finland

^173^Clinical Neurosciences, Neurology, University of Helsinki, Helsinki, Finland

^174^Department of Neurology, University of Washington, Seattle, WA, USA

^175^Albrecht Kossel Institute, University Clinic of Rostock, Rostock, Germany

^176^Clinical Trial Service Unit and Epidemiological Studies Unit, Nuffield Department of Population Health, University of Oxford, Oxford, UK

^177^Department of Genetics, Perelman School of Medicine, University of Pennsylvania, PA, USA

^178^Faculty of Medicine, University of Iceland, Reykjavik, Iceland

^179^Departments of Neurology and Public Health Sciences, University of Virginia School of Medicine, Charlottesville, VA, USA

^180^Department of Neurology, Boston University School of Medicine, Boston, MA, USA

^181^Human Genetics Center, University of Texas Health Science Center at Houston, Houston, TX, USA

^182^Center for Genomic Medicine, Kyoto University Graduate School of Medicine, Kyoto, Japan

^183^Munich Cluster for Systems Neurology (SyNergy), Munich, Germany

^184^German Center for Neurodegenerative Diseases (DZNE), Munich, Germany

^185^Boston University School of Medicine, Boston, MA, USA

^186^University of Kentucky College of Public Health, Lexington, KY, USA

^187^University of Newcastle and Hunter Medical Research Institute, New Lambton, Australia

^188^Univ. Montpellier, Inserm, U1061, Montpellier, France

^189^Centre for Research in Environmental Epidemiology, Barcelona, Spain

^190^Department of Neurology, Università degli Studi di Perugia, Umbria, Italy

^191^Department of Medicine, University of Maryland School of Medicine, Baltimore, MD, USA

^192^Broad Institute, Cambridge, MA, USA

^193^Univ. Bordeaux, Inserm, Bordeaux Population Health Research Center, UMR 1219, Bordeaux, France

^194^Bordeaux University Hospital, Department of Neurology, Memory Clinic, Bordeaux, France

^195^Neurovascular Research Laboratory. Vall d’Hebron Institut of Research, Neurology and Medicine Departments-Universitat Autònoma de Barcelona. Vall d’Hebrón Hospital, Barcelona, Spain

^196^University Medicine Greifswald, Department of Internal Medicine B, Greifswald, Germany

^197^DZHK, Greifswald, Germany

^198^Robertson Center for Biostatistics, University of Glasgow, Glasgow, UK

^199^Hero DMC Heart Institute, Dayanand Medical College & Hospital, Ludhiana, India

^200^Atherosclerosis Research Unit, Department of Medicine Solna, Karolinska Institutet, Stockholm, Sweden

^201^Karolinska Institutet, Stockholm, Sweden

^202^Division of Emergency Medicine, and Department of Neurology, Washington University School of Medicine, St. Louis, MO, USA

^203^Tohoku Medical Megabank Organization, Sendai, Japan

^204^Department of Psychiatry, Washington University School of Medicine, St. Louis, MO, USA

^205^Department of Public Health and Caring Sciences / Geriatrics, Uppsala University, Uppsala, Sweden

^206^Epidemiology and Prevention Group, Center for Public Health Sciences, National Cancer Center, Tokyo, Japan

^207^Department of Internal Medicine and the Center for Clinical and Translational Science, The Ohio State University, Columbus, OH, USA

^208^Institute of Neuroscience and Physiology, the Sahlgrenska Academy at University of Gothenburg, Goteborg, Sweden

^209^Department of Basic and Clinical Neurosciences, King’s College London, London, UK

^210^Department of Health Care Administration and Management, Graduate School of Medical Sciences, Kyushu University, Japan

^211^Department of Medicine and Clinical Science, Graduate School of Medical Sciences, Kyushu University, Japan

^212^Landspitali National University Hospital, Departments of Neurology & Radiology, Reykjavik, Iceland

^213^Department of Neurology, Heidelberg University Hospital, Germany

^214^Department of Neurology, Erasmus University Medical Center

^215^Hospital Universitari Mutua Terrassa, Terrassa (Barcelona), Spain

^216^Albert Einstein College of Medicine, Montefiore Medical Center, New York, NY, USA

^217^John Hunter Hospital, Hunter Medical Research Institute and University of Newcastle, Newcastle, NSW, Australia

^218^Centre for Prevention of Stroke and Dementia, Nuffield Department of Clinical Neurosciences, University of Oxford, UK

^219^Department of Medical Sciences, Uppsala University, Uppsala, Sweden

^220^Genetic and Genomic Epidemiology Unit, Wellcome Trust Centre for Human Genetics, University of Oxford, Oxford, UK

^221^The Wellcome Trust Centre for Human Genetics, Oxford, UK

^222^Beth Israel Deaconess Medical Center, Boston, MA, USA

^223^Wake Forest School of Medicine, Wake Forest, NC, USA

^224^Department of Neurology, University of Pittsburgh, Pittsburgh, PA, USA

^225^BioBank Japan, Laboratory of Clinical Sequencing, Department of Computational biology and medical Sciences, Graduate school of Frontier Sciences, The University of Tokyo, Tokyo, Japan

^226^Neurovascular Research Laboratory, Vall d’Hebron Institut of Research, Neurology and Medicine Departments-Universitat Autònoma de Barcelona. Vall d’Hebrón Hospital, Barcelona, Spain

^227^Department of Biostatistics, University of Liverpool, Liverpool, UK

^228^Wellcome Trust Centre for Human Genetics, University of Oxford, Oxford, UK

^229^Institute of Genetic Epidemiology, Helmholtz Zentrum München - German Research Center for Environmental Health, Neuherberg, Germany

^230^Department of Medicine I, Ludwig-Maximilians-Universität, Munich, Germany

^231^DZHK (German Centre for Cardiovascular Research), partner site Munich Heart Alliance, Munich, Germany

^232^Department of Cerebrovascular Diseases, Fondazione IRCCS Istituto Neurologico “Carlo Besta”, Milano, Italy

^233^Karolinska Institutet, MEB, Stockholm, Sweden

^234^University of Tartu, Estonian Genome Center, Tartu, Estonia, Tartu, Estonia

^235^Department of Clinical and Experimental Sciences, Neurology Clinic, University of Brescia, Italy

^236^Translational Genomics Unit, Department of Oncology, IRCCS Istituto di Ricerche Farmacologiche Mario Negri, Milano, Italy

^237^Department of Genetics, Microbiology and Statistics, University of Barcelona, Barcelona, Spain

^238^Psychiatric Genetics Unit, Group of Psychiatry, Mental Health and Addictions, Vall d’Hebron Research Institute (VHIR), Universitat Autònoma de Barcelona, Biomedical Network Research Centre on Mental Health (CIBERSAM), Barcelona, Spain

^239^Department of Neurology, IMIM-Hospital del Mar, and Universitat Autònoma de Barcelona, Spain

^240^IMIM (Hospital del Mar Medical Research Institute), Barcelona, Spain

^241^National Institute for Health Research Comprehensive Biomedical Research Centre, Guy’s & St. Thomas’ NHS Foundation Trust and King’s College London, London, UK

^242^Division of Health and Social Care Research, King’s College London, London, UK

^243^FIMM-Institute for Molecular Medicine Finland, Helsinki, Finland

^244^THL-National Institute for Health and Welfare, Helsinki, Finland

^245^Iwate Tohoku Medical Megabank Organization, Iwate Medical University, Iwate, Japan

^246^BHF Glasgow Cardiovascular Research Centre, Faculty of Medicine, Glasgow, UK

^247^deCODE Genetics/Amgen, Inc., Reykjavik, Iceland

^248^Icelandic Heart Association, Reykjavik, Iceland

^249^Institute of Biomedicine, the Sahlgrenska Academy at University of Gothenburg, Goteborg, Sweden

^250^Department of Epidemiology, University of Maryland School of Medicine, Baltimore, MD, USA

^251^Institute of Cardiovascular and Medical Sciences, Faculty of Medicine, University of Glasgow, Glasgow, UK

^252^Chair of Genetic Epidemiology, IBE, Faculty of Medicine, LMU Munich, Germany

^253^Division of Epidemiology and Prevention, Aichi Cancer Center Research Institute, Nagoya, Japan

^254^Department of Epidemiology, Nagoya University Graduate School of Medicine, Nagoya, Japan

^255^University Medicine Greifswald, Institute for Community Medicine, SHIP-KEF, Greifswald, Germany

^256^Department of Neurology, Caen University Hospital, Caen, France

^257^University of Caen Normandy, Caen, France

^258^Department of Internal Medicine, Erasmus University Medical Center, Rotterdam, Netherlands

^259^Landspitali University Hospital, Reykjavik, Iceland

^260^Survey Research Center, University of Michigan, Ann Arbor, MI, USA

^261^University of Virginia Department of Neurology, Charlottesville, VA, USA

